# HELMET: A Hybrid Machine Learning Framework for Real-Time Prediction of Edema Trajectory in Large Middle Cerebral Artery Stroke

**DOI:** 10.1101/2024.11.13.24317229

**Authors:** Ethan Phillips, Odhran O’Donoghue, Yumeng Zhang, Panos Tsimpos, Leigh Ann Mallinger, Stefanos Chatzidakis, Jack Pohlmann, Yili Du, Ivy Kim, Jonathan Song, Benjamin Brush, Stelios Smirnakis, Charlene J Ong, Agni Orfanoudaki

**Author notes:** Co-lead study authors (Charlene J Ong, Agni Orfanoudaki).

## Abstract

Malignant cerebral edema occurs when brain swelling displaces and compresses vital midline structures within the first week of a large middle cerebral artery stroke. Early interventions such as hyperosmolar therapy or surgical decompression may reverse secondary injury but must be administered judiciously. To optimize treatment and reduce secondary damage, clinicians need strategies to frequently and quantitatively assess the trajectory of edema using updated, relevant information. However, existing risk assessment tools are limited by the absence of structured records capturing the evolution of edema and typically estimate risk at a single time point early in the admission, therefore failing to account for changes in variables over the following hours or days. To address this, we developed and validated dynamic machine learning models capable of accurately predicting the severity of midline structure displacement, an established indicator of malignant edema, in real-time. Our models can provide updated estimations as frequently as every hour, using data from structured time-varying patient records, radiographic text, and human-curated neurological characteristics. Our work resulted in two novel multi-class classification models, collectively named Hybrid Ensemble Learning Models for Edema Trajectory (HELMET), predicting the progression of midline shift over 8-hour (HELMET-8) and 24-hour windows (HELMET-24), respectively. HELMET combines transformer-based large language models with supervised ensemble learning, demonstrating the value of merging human expertise and multimodal health records in developing clinical risk scores. Both models were trained on a retrospective cohort of 15,696 observations from 623 patients hospitalized with large middle cerebral artery ischemic stroke and were externally validated using 3,713 observations from 60 patients at a separate hospital system. Our HELMET models are accurate and generalize effectively to diverse populations, achieving a cross-validated mean area under the receiver operating characteristic score of 96.6% in the derivation cohort and 92.5% in the external validation cohort. Moreover, our approach provides a framework for developing hybrid risk prediction models that integrate both human-extracted and algorithm-derived multi-modal inputs. Our work enables accurate estimation of complex, dynamic, and highly specific clinical targets, such as midline shift, in real-time, even when relevant structured information is limited in electronic health record databases.

## 1 Introduction

Large middle cerebral artery (MCA) infarction is a potentially lethal form of stroke, occurring in between 18% and 31% of ischemic stroke cases involving MCA occlusion [1]. A major driver of poor stroke outcomes is malignant cerebral edema, which can result in a 40-80% risk of neurological deterioration and death [1]. Specifically, space-occupying malignant edema displaces and compresses the surrounding brain tissue, causing further damage referred to as mass effect [1–6].

Early recognition of evolving cerebral edema is imperative as treatments, such as surgical decompression, can reduce the risk of mortality from 80% to 20% in eligible patients [7, 8]. Pharmaceutical strategies, such as hyperosmolar therapy, are also widely employed by intensivists in efforts to treat worsening or life-threatening edema [9, 10]. However, the course of malignant edema can be unpredictable, varying rapidly over mere hours or more slowly over multiple days. For this reason, stroke patients at risk of malignant edema are often monitored in intensive care unit settings to closely watch for signs of neurological deterioration.

In practice, clinicians use a variety of dynamic information available to them to monitor the risk of edema for individual patients at specific time points, including physical exam assessments, laboratory data, vital signs, and neuroimaging results [9]. Neuroimaging techniques, often in the form of computed tomography (CT) and magnetic resonance imaging, are particularly important for determining the extent of edema as they provide quantitative evidence of edema progression [8, 11]. A common clinical marker is the midline shift (MLS) of the septum pellucidum, measured in millimeters of displacement from the cerebral midline [8, 11]. MLS is a measurable, quantifiable, and clinically relevant indicator of worsening mass effect, used to standardize communication for edema severity, and thus becomes a key determinant of stroke patient treatment and management decisions [8, 12, 13]. Prior studies have shown that MLS greater than 5mm within the first two days is associated with neurological deterioration and early mortality [13]. More recently, MLS as low as 3mm has been shown to be associated with worse long-term outcomes [12].

Current strategies for monitoring mass effect and other secondary injuries primarily rely on physical examination and confirmatory imaging. However, guidelines recognize that the clinical practice of detecting arousal depression due to mass effect is often inadequate, as it may only become evident after a significant secondary injury has already occurred [1]. This challenge is compounded in patients whose mental status is already depressed from other factors, including the initial injury, medications, fever, or toxic-metabolic abnormalities. Therefore, in the absence of continuous neuroimaging, early signs of deterioration may go unnoticed. Near-continuous CT imaging is impractical due to constraints on CT availability, risks associated with patient transport, and concerns regarding radiation exposure [14, 15]. Imaging in many clinical settings is often limited to once every 24 hours, or prompted only by clear signs of clinical deterioration. As a result, current guidelines do not specify the optimal frequency for surveillance CT imaging to monitor edema progression, leaving decisions to clinician discretion and contributing to variability in practice and quality of care. [9, 13, 16].

Data-driven models for edema risk assessment could lead to more personalized and accurate screening policies. However, existing models predicting cerebral edema risk [17–20] rely on structured and curated data collected early in hospitalization to forecast late clinical outcomes, such as death or the need for surgical decompression after medical interventions have taken place (see also Table 1 in the Supplementary Information). Relevant predictors identified by these models include approximations of higher infarct volume (such as the national institutes of health stroke scale (NIHSS)), baseline laboratory values, and severe early mass effect [17, 19–21]. Limited research exists on effectively integrating dynamic changes in these variables into standardized risk assessments, which is critical for providers making real-time decisions across iterative time points. In addition to limited external validation and relatively small sample sizes[17–19], currently available static models often lack utility for personalized decision-making as new information becomes available, and are therefore not frequently consulted by physicians in practice. This is a significant limitation, particularly for patients where treatment decisions, including surgical decompression, are made outside the time frame for which there is high-quality evidence of efficacy (<48 hours) [22].

**Table 1:**
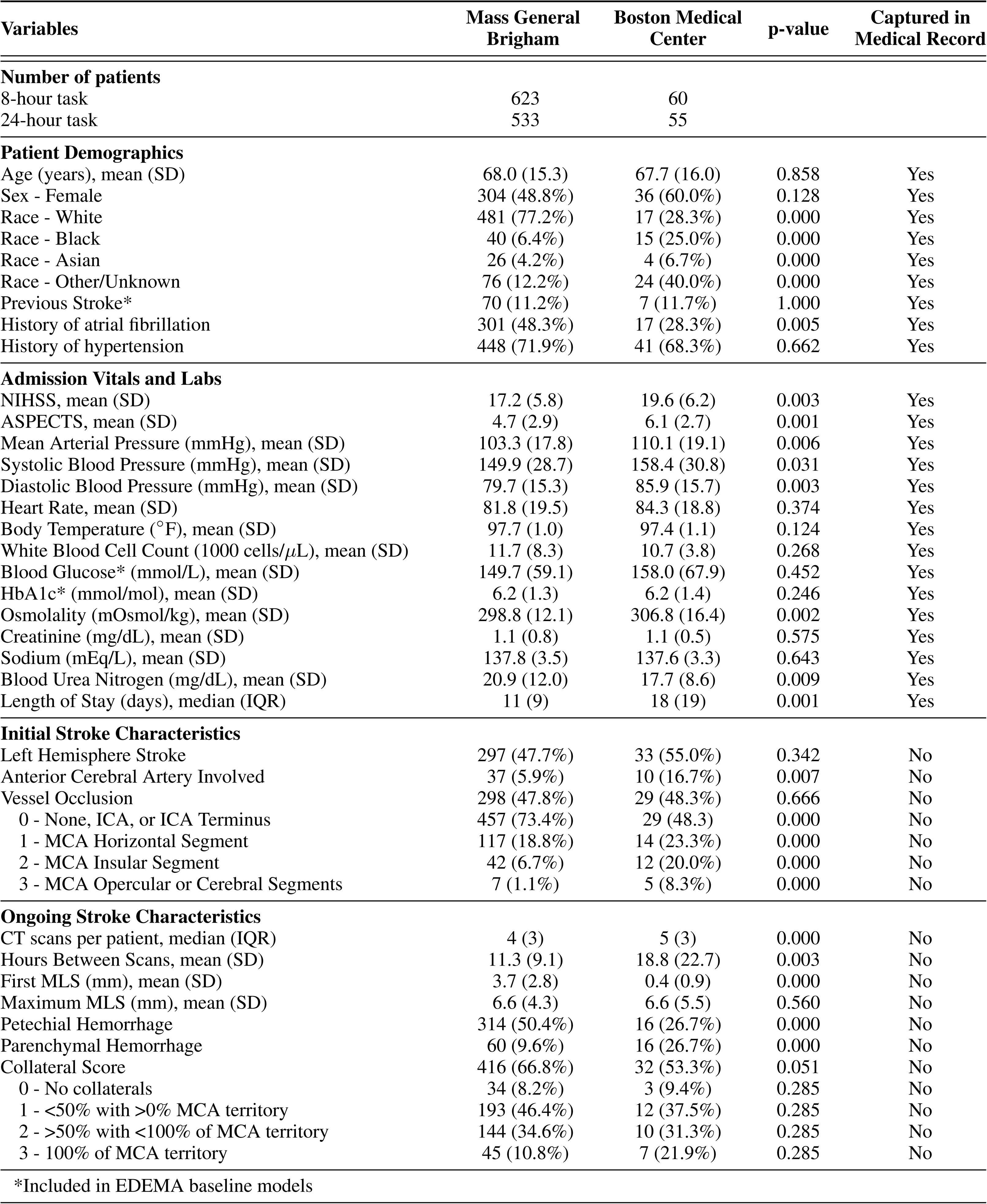
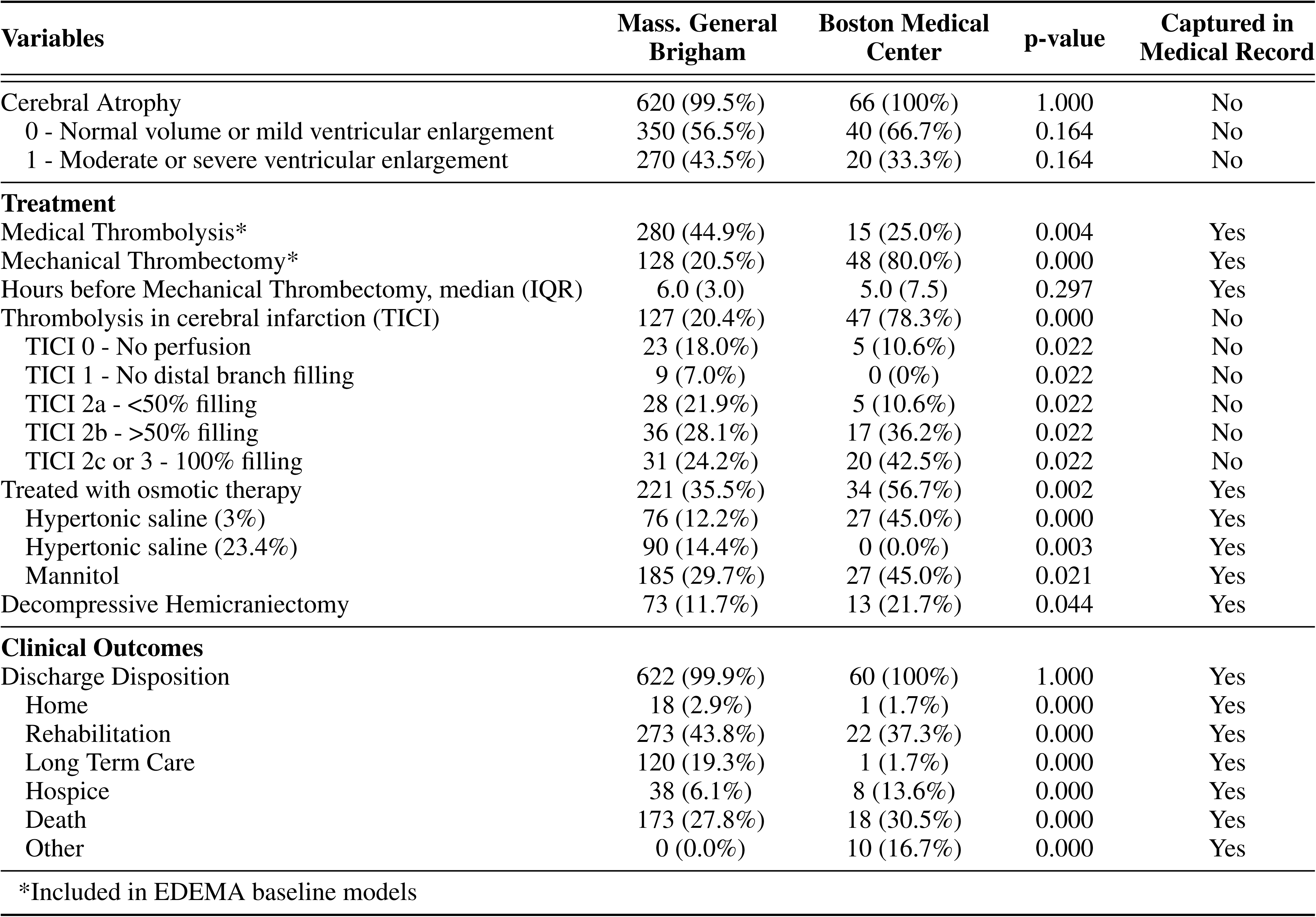
Basic patient characteristics across the derivation and external validation datasets. All descriptive statistics were calculated using the 8-hour task patient cohorts. Continuous variables are reported as the mean (standard deviation) and used either a two-sample t-test or the Mann-Whitney U test for significance testing, depending on normality. Categorical or binary variables are reported as the patient count (proportion) and used the *χ*^2^ test for significance testing.

| Variables | Mass General Brigham | Boston Medical Center | p-value | Captured in Medical Record |
| --- | --- | --- | --- | --- |
| **Number of patients** |  |  |  |  |
| 8-hour task | 623 | 60 |  |  |
| 24-hour task | 533 | 55 |  |  |
| **Patient Demographics** |  |  |  |  |
| Age (years), mean (SD) | 68.0 (15.3) | 67.7 (16.0) | 0.858 | Yes |
| Sex - Female | 304 (48.8%) | 36 (60.0%) | 0.128 | Yes |
| Race - White | 481 (77.2%) | 17 (28.3%) | 0.000 | Yes |
| Race - Black | 40 (6.4%) | 15 (25.0%) | 0.000 | Yes |
| Race - Asian | 26 (4.2%) | 4 (6.7%) | 0.000 | Yes |
| Race - Other/Unknown | 76 (12.2%) | 24 (40.0%) | 0.000 | Yes |
| Previous Stroke* | 70 (11.2%) | 7 (11.7%) | 1.000 | Yes |
| History of atrial fibrillation | 301 (48.3%) | 17 (28.3%) | 0.005 | Yes |
| History of hypertension | 448 (71.9%) | 41 (68.3%) | 0.662 | Yes |
| **Admission Vitals and Labs** |  |  |  |  |
| NIHSS, mean (SD) | 17.2 (5.8) | 19.6 (6.2) | 0.003 | Yes |
| ASPECTS, mean (SD) | 4.7 (2.9) | 6.1 (2.7) | 0.001 | Yes |
| Mean Arterial Pressure (mmHg), mean (SD) | 103.3 (17.8) | 110.1 (19.1) | 0.006 | Yes |
| Systolic Blood Pressure (mmHg), mean (SD) | 149.9 (28.7) | 158.4 (30.8) | 0.031 | Yes |
| Diastolic Blood Pressure (mmHg), mean (SD) | 79.7 (15.3) | 85.9 (15.7) | 0.003 | Yes |
| Heart Rate, mean (SD) | 81.8 (19.5) | 84.3 (18.8) | 0.374 | Yes |
| Body Temperature (°F), mean (SD) | 97.7 (1.0) | 97.4 (1.1) | 0.124 | Yes |
| White Blood Cell Count (1000 cells/μL), mean (SD) | 11.7 (8.3) | 10.7 (3.8) | 0.268 | Yes |
| Blood Glucose* (mmol/L), mean (SD) | 149.7 (59.1) | 158.0 (67.9) | 0.452 | Yes |
| HbA1c* (mmol/mol), mean (SD) | 6.2 (1.3) | 6.2 (1.4) | 0.246 | Yes |
| Osmolality (mOsmol/kg), mean (SD) | 298.8 (12.1) | 306.8 (16.4) | 0.002 | Yes |
| Creatinine (mg/dL), mean (SD) | 1.1 (0.8) | 1.1 (0.5) | 0.575 | Yes |
| Sodium (mEq/L), mean (SD) | 137.8 (3.5) | 137.6 (3.3) | 0.643 | Yes |
| Blood Urea Nitrogen (mg/dL), mean (SD) | 20.9 (12.0) | 17.7 (8.6) | 0.009 | Yes |
| Length of Stay (days), median (IQR) | 11 (9) | 18 (19) | 0.001 | Yes |
| **Initial Stroke Characteristics** |  |  |  |  |
| Left Hemisphere Stroke | 297 (47.7%) | 33 (55.0%) | 0.342 | No |
| Anterior Cerebral Artery Involved | 37 (5.9%) | 10 (16.7%) | 0.007 | No |
| Vessel Occlusion | 298 (47.8%) | 29 (48.3%) | 0.666 | No |
| 0 - None, ICA, or ICA Terminus | 457 (73.4%) | 29 (48.3) | 0.000 | No |
| 1 - MCA Horizontal Segment | 117 (18.8%) | 14 (23.3%) | 0.000 | No |
| 2 - MCA Insular Segment | 42 (6.7%) | 12 (20.0%) | 0.000 | No |
| 3 - MCA Opercular or Cerebral Segments | 7 (1.1%) | 5 (8.3%) | 0.000 | No |
| **Ongoing Stroke Characteristics** |  |  |  |  |
| CT scans per patient, median (IQR) | 4 (3) | 5 (3) | 0.000 | No |
| Hours Between Scans, mean (SD) | 11.3 (9.1) | 18.8 (22.7) | 0.003 | No |
| First MLS (mm), mean (SD) | 3.7 (2.8) | 0.4 (0.9) | 0.000 | No |
| Maximum MLS (mm), mean (SD) | 6.6 (4.3) | 6.6 (5.5) | 0.560 | No |
| Petechial Hemorrhage | 314 (50.4%) | 16 (26.7%) | 0.000 | No |
| Parenchymal Hemorrhage | 60 (9.6%) | 16 (26.7%) | 0.000 | No |
| Collateral Score | 416 (66.8%) | 32 (53.3%) | 0.051 | No |
| 0 - No collaterals | 34 (8.2%) | 3 (9.4%) | 0.285 | No |
| 1 - <50% with >0% MCA territory | 193 (46.4%) | 12 (37.5%) | 0.285 | No |
| 2 - >50% with <100% of MCA territory | 144 (34.6%) | 10 (31.3%) | 0.285 | No |
| 3 - 100% of MCA territory | 45 (10.8%) | 7 (21.9%) | 0.285 | No |

\*Included in EDEMA baseline models

Table 1 – continued from previous page
| Variables | Mass. General Brigham | Boston Medical Center | p-value | Captured in Medical Record |
| --- | --- | --- | --- | --- |
| Cerebral Atrophy | 620 (99.5%) | 66 (100%) | 1.000 | No |
| 0 - Normal volume or mild ventricular enlargement | 350 (56.5%) | 40 (66.7%) | 0.164 | No |
| 1 - Moderate or severe ventricular enlargement | 270 (43.5%) | 20 (33.3%) | 0.164 | No |
| **Treatment** |  |  |  |  |
| Medical Thrombolysis* | 280 (44.9%) | 15 (25.0%) | 0.004 | Yes |
| Mechanical Thrombectomy* | 128 (20.5%) | 48 (80.0%) | 0.000 | Yes |
| Hours before Mechanical Thrombectomy, median (IQR) | 6.0 (3.0) | 5.0 (7.5) | 0.297 | Yes |
| Thrombolysis in cerebral infarction (TICI) | 127 (20.4%) | 47 (78.3%) | 0.000 | No |
| TICI 0 - No perfusion | 23 (18.0%) | 5 (10.6%) | 0.022 | No |
| TICI 1 - No distal branch filling | 9 (7.0%) | 0 (0%) | 0.022 | No |
| TICI 2a - <50% filling | 28 (21.9%) | 5 (10.6%) | 0.022 | No |
| TICI 2b - >50% filling | 36 (28.1%) | 17 (36.2%) | 0.022 | No |
| TICI 2c or 3 - 100% filling | 31 (24.2%) | 20 (42.5%) | 0.022 | No |
| Treated with osmotic therapy | 221 (35.5%) | 34 (56.7%) | 0.002 | Yes |
| Hypertonic saline (3%) | 76 (12.2%) | 27 (45.0%) | 0.000 | Yes |
| Hypertonic saline (23.4%) | 90 (14.4%) | 0 (0.0%) | 0.003 | Yes |
| Mannitol | 185 (29.7%) | 27 (45.0%) | 0.021 | Yes |
| Decompressive Hemicraniectomy | 73 (11.7%) | 13 (21.7%) | 0.044 | Yes |
| **Clinical Outcomes** |  |  |  |  |
| Discharge Disposition | 622 (99.9%) | 60 (100%) | 1.000 | Yes |
| Home | 18 (2.9%) | 1 (1.7%) | 0.000 | Yes |
| Rehabilitation | 273 (43.8%) | 22 (37.3%) | 0.000 | Yes |
| Long Term Care | 120 (19.3%) | 1 (1.7%) | 0.000 | Yes |
| Hospice | 38 (6.1%) | 8 (13.6%) | 0.000 | Yes |
| Death | 173 (27.8%) | 18 (30.5%) | 0.000 | Yes |
| Other | 0 (0.0%) | 10 (16.7%) | 0.000 | Yes |

\*Included in EDEMA baseline models

For these reasons, improved methods to estimate cerebral edema trajectory are needed to leverage the rich sources of structured and unstructured data currently available in electronic health records. Readily-available, dynamic, and accurate MLS severity estimation tools could assist medical providers in making more timely treatment decisions, improving operational and clinical outcomes, and delivering more personalized care. Recent efforts to dynamically identify other important hospitalization events, including sepsis and non-neurological clinical deterioration, reinforce the need and potential utility of dynamic forecasting in neurocritical care settings [23, 24].

To this end, we curated a retrospective, multi-modal, multi-institutional dataset comprised of both static and time-varying variables from electronic health records, radiographic report texts, and expert-labeled neuroanatomic features derived from radiographic images [21, 25, 26]. This multi-modal approach builds on prior research showing that integrating data of different types from multiple sources into a single model leads to improved predictive performance over models which used just one data type [27]. Using this dataset, we developed and externally validated the Hybrid Ensemble Learning Models for Edema Trajectory (HELMET) to predict worsening MLS class (0 mm, 0–3 mm, 3–8 mm, and >8 mm) within 8-hour (HELMET-8) and 24-hour (HELMET-24) windows. We employed a combination of machine learning techniques, including large language models for the interpretation of the raw radiographic texts and an ensemble learning algorithm for downstream final predictions from structured data inputs. HELMET provides a paradigm for the development and validation of dynamic prediction scores for complex and volatile targets that are not routinely captured within structured electronic health records.

Our aim is to complement existing non-temporal cerebral edema prediction models and build on their successes by providing more granular data to inform imaging and treatment decisions at critical moments in a generalizable way. By incorporating data from two distinct academic medical centers with differing patient pool demographics, our study demonstrates the generalizability of HELMET in diverse patient populations [28]. Our work can assist in prompting earlier life-saving interventions and more efficient resource use by making edema progression predictions accessible to clinical teams at dynamic time horizons. We believe our findings represent the first step toward developing policies that alert the clinical team to evolving secondary injury and aid in the appropriate use of diagnostic testing.

## 2 Results

### 2.1 Study Population

The derivation cohort consists of 623 patients with acute MCA ischemic stroke, affecting at least half of the MCA territory, who were retrospectively identified from admissions to Massachusetts General Hospital and Brigham and Women’s Hospital—core institutions of the Mass General Brigham healthcare system in Boston, Massachusetts—between January 2006 and July 2021. We also leveraged a prospective external validation cohort of 60 patients with acute MCA ischemic stroke, affecting at least half of the MCA territory, admitted to Boston Medical Center between May 2019 and November 2023, drawn from an existing available dataset originally compiled for pupillometry research. The latter constitutes the largest safety-net hospital in New England with a different racial and socioeconomic patient population makeup than Mass General Brigham. The derivation cohort was used to train the HELMET models, while the Boston Medical Center cohort, selected for its diverse patient population, served as an independent dataset for external validation. The full patient inclusion diagrams for both cohorts are shown in Figure 1. Information on the exclusion criteria can be found in Section 4.1.

**Figure 1:**
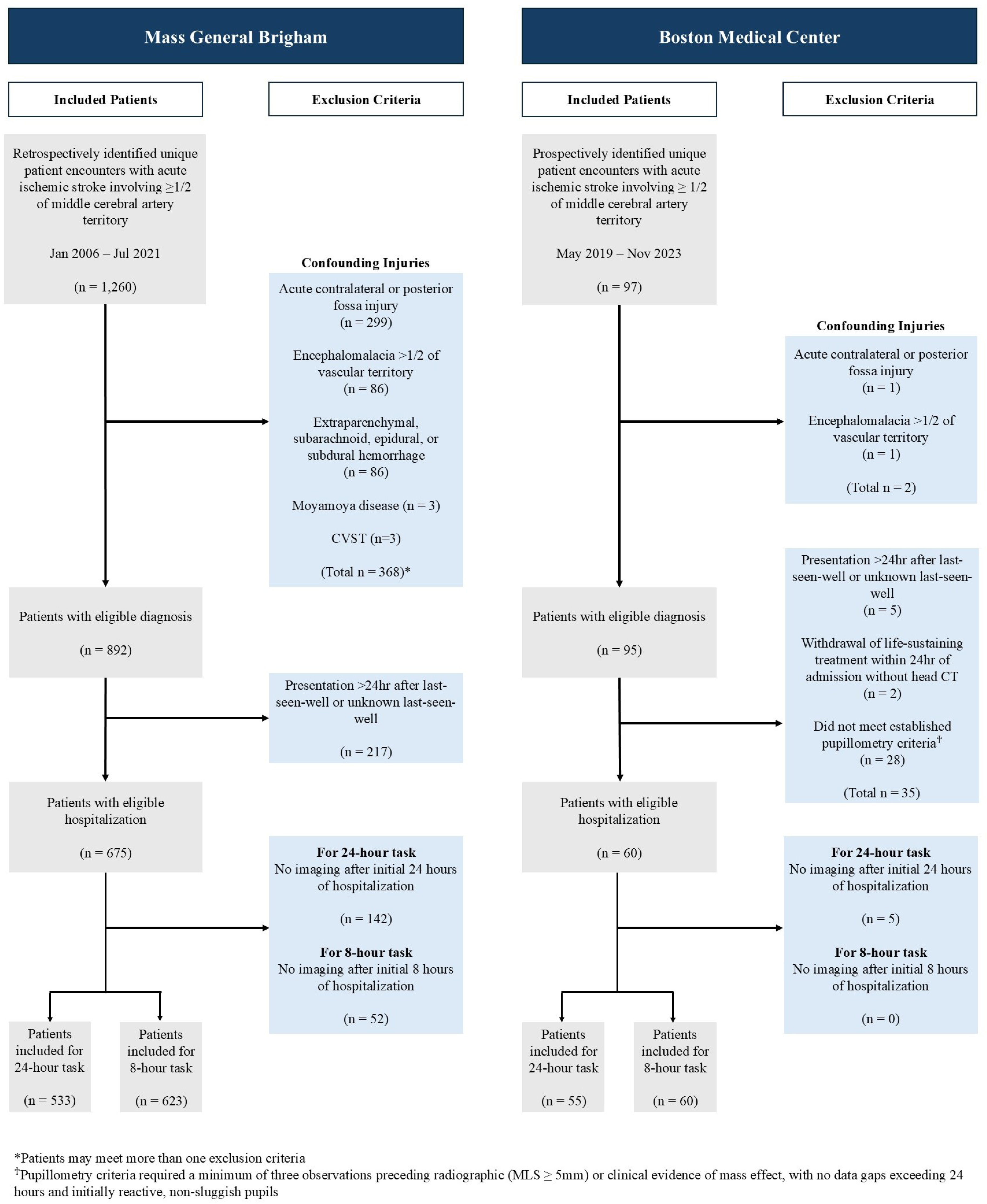
Patient inclusion for the derivation cohort and the external validation cohort. After applying exclusion criteria and removing patients with insufficient data, 623 patients were included in the derivation cohort dataset^†^ for the 8-hour task, and 60 patients were included in the external validation cohort dataset for the 8-hour task. For the 24-hour task, 533 patients were included in the derivation cohort and 55 patients were included in the external validation cohort.

Table 1 summarizes the characteristics of both patient cohorts. For the Mass General Brigham cohort, the average age was 68.0 years, and 48.8% of the patients were female. In the Boston Medical Center cohort, the average age was 67.7 years, and 60.0% of the patients were female. A statistically significant difference in racial composition was found between the two cohorts, with the Boston Medical Center cohort having a higher proportion of non-White patients (*p*-value*<* 0.001). The Boston Medical Center cohort also had worse average stroke severity indicators at admission (NIHSS and the Alberta Stroke Programme Early CT Score (ASPECTS)) by a small but statistically significant margin (*p*-values*<* 0.005). We focused on predictions in the first seven days following patient presentation to the hospital and truncated data beyond this point. For full details of our inclusion criteria and data transformations, please see the Methods section.

To derive dynamic prediction models of edema trajectory, longitudinal patient data were transformed to per-hour *observations* for each patient over the course of their hospitalization (described in Section 4.2). For the Mass General Brigham dataset (derivation cohort), data transformations resulted in 8,515 observations (patient-hours) for the 8-hour prediction task and 15,696 observations for the 24-hour prediction task. For the Boston Medical Center dataset (external validation cohort), transformations resulted in 1,891 observations for the 8-hour prediction task and 3,713 observations for the 24-hour prediction task. Total observations for each outcome class and horizon prediction task are reported in Table 2. The breakdown of maximum MLS class reached by each patient is presented in Table 2 of the Supplementary Information.

**Table 2:** Number of observations^†^ per task and class of prediction target.

| Data set | Task | Maximum MLS class |  |  |  |
| --- | --- | --- | --- | --- | --- |
|  |  | 0mm | 0-3mm | 3-8mm | >8mm |
| Mass General Brigham<br>(derivation cohort) | 8-hour | 2012 | 1568 | 3405 | 1530 |
|  | 24-hour | 2638 | 2713 | 6867 | 3478 |
| Boston Medical Center<br>(validation cohort) | 8-hour | 523 | 309 | 620 | 439 |
|  | 24-hour | 873 | 696 | 1223 | 921 |

<sup>†</sup>Observations are transformed hourly sets of data for a given patient. Each patient may account for up to 8 observations per scan on the 8-hour task, and up to 24 observations per scan on the 24-hour task.

### 2.2 Model Performance

The HELMET models were trained using randomized five-fold bootstrapped partitions of the derivation cohort patients, and evaluated on the remaining test set patients from the Massachusetts General Brigham derivation cohort as well as the full external validation cohort from Boston Medical Center. Two separate models were trained to predict MLS severity on 24-hour (HELMET-24) and 8-hour (HELMET-8) horizons, respectively. The HELMET models were retrospectively evaluated in head-to-head comparisons against multinomial regression models trained separately in the derivation and external validation cohorts^1^ using the same input features as the preexisting EDEMA score, originally developed to predict potentially life-threatening malignant edema [17]. Figure 2 provides an illustration of the dataset curation, feature extraction, and model derivation process.

**Figure 2:**
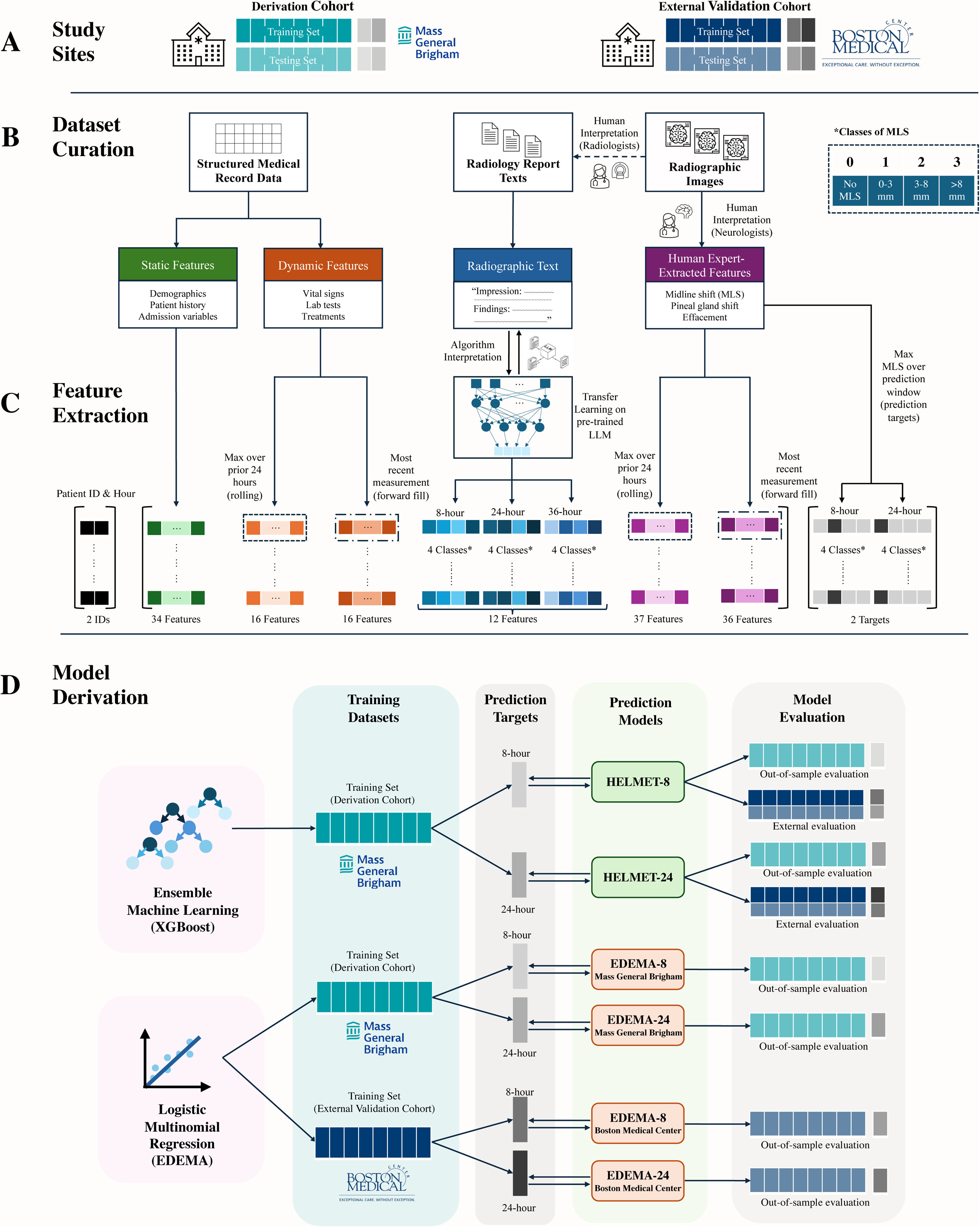
Methods process flow diagram. Across the panels, we show the study site cohorts (A); multimodal dataset construction using static, time-varying, and text-based variables (B); transformations applied to the data to generate observations (C); and subsequent model training (D).

Each model’s performance was assessed using both the original dataset encompassing all observations (overall metrics) as well as a filtered dataset including only observations where the patient’s current MLS state differed from their future target class within the predictive window (filtered metrics), to further analyze model performance on observations capturing clinically relevant transitions in MLS state. Figure 3 illustrates in radar plots the overall and filtered performance of each model for each task and cohort. The five radial axes represent the performance metrics of sensitivity, specificity, accuracy, area under the precision-recall curve (AUPRC), and area under the receiver operator curve (AUROC). Across all measures, higher values are plotted further from the center, indicating superior predictive performance.^2^

**Figure 3:**
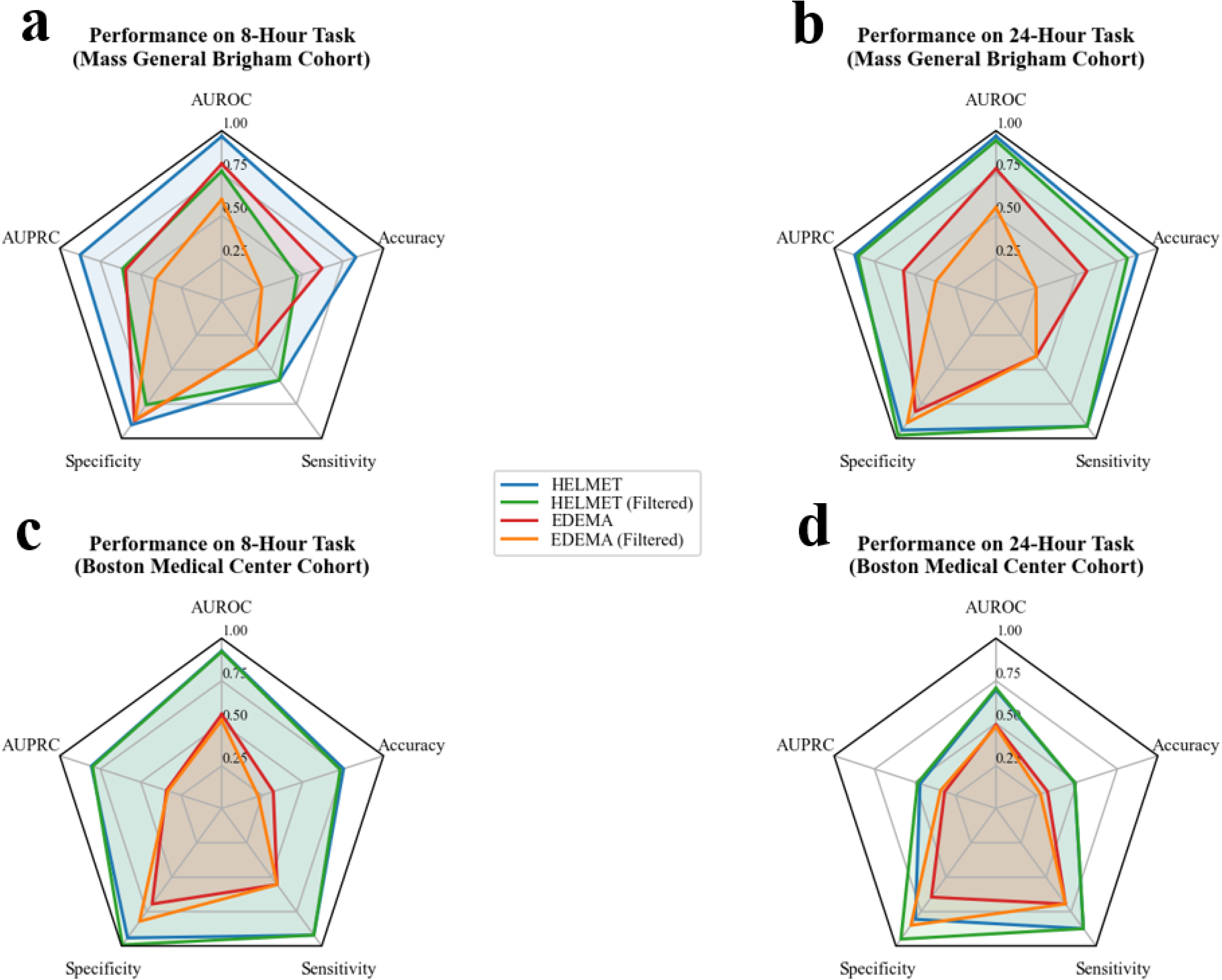
Radar plot comparison of the HELMET models and baseline EDEMA models performance across all cohorts and prediction tasks. Each radial axis represents a distinct performance metric, with higher performance values further from center. Polyhedrons with greater total area on the plots show higher-performing models, and smaller polyhedrons show lower-performing models. Blue lines represent the overall performance for the HELMET models, green lines represent the filtered performance for the HELMET models, red lines represent the overall performance for the baseline EDEMA models, and orange lines represent the filtered performance for the baseline EDEMA models. Specifically, we summarize model performance across tasks and cohorts in the following panels: a) derivation cohort for the 8-hour prediction task; b) derivation cohort for the 24-hour prediction task; c) external validation cohort for the 8-hour prediction task; d) external validation cohort for the 24-hour task.

In the out-of-sample evaluation of the derivation cohort, the large language model-enhanced HELMET-24 model achieved a mean AUROC score of 96.7%, sensitivity of 91.2%, and specificity of 94.0%. These scores outperform the EDEMA-24 baseline by 19.0 percentage points in AUROC, 50.9 percentage points in sensitivity, and 13.4 percentage points in specificity. On the 8-hour prediction task, HELMET-8 resulted in a mean AUROC of 96.6%, sensitivity of 82.9%, and specificity of 92.7%, outperforming the EDEMA-8 model by 16.1, 48.6, and 5.6 percentage points across the three metrics, respectively. In the dataset filtered for changes in MLS severity class, both HELMET-24 and HELMET-8 outperformed baseline EDEMA models. HELMET-24 achieved a mean filtered AUROC of 94.2% compared to 55.5% for the baseline, while HELMET-8 achieved 76.1% compared to baseline performance of 59.7%.

To assess the generalizability of our models, we also evaluated the HELMET models on the entirety of the external validation cohort. We observed that the HELMET models consistently outperform the EDEMA baseline models by a significant margin across both sites and both tasks. HELMET-24 achieved a mean overall AUROC of 69.7% and a mean filtered AUROC of 70.7% on the 24-hour task, outperforming the EDEMA-24 model by 20.7 percentage points on overall AUROC and 22.7 percentage points on filtered AUROC. On the 8-hour task, HELMET-8 achieved an overall AUROC of 92.5% and a filtered AUROC of 92.1%, outperforming EDEMA-8 by 37.2 and 40.3 percentage points respectively.

The complete table of performance metrics, including respective 95% confidence intervals, is presented in Table 9 of the Supplementary Information. Receiver operator characteristic and precision-recall curves are shown in Figures 1 and 2 in the Supplementary Information.

### 2.3 Model Feature Interpretation

To determine the relative importance of contributing features to our models, we applied the Shapley Additive Explanation framework on each of the four prediction MLS classes of HELMET-24 and HELMET-8 (see also Section 4.4.2). Figure 4 illustrates the composition of the 20 most important features, ranked by Shapley Additive Explanation values, in the HELMET models for the 24-hour and 8-hour prediction tasks, categorized by the most recent prior MLS class. The Shapley analysis allowed us to gain insight into the interplay of the different data sources comprising the HELMET predictions. Specifically, our results indicate that the large language model predictions on radiological report texts are highly important across both the 8-hour and 24-hour prediction task models. Human-extracted features by neurology experts, which may not always be present in the dictated radiology reports, and their associated times make up the second-largest category of high-impact features. We also observe that while dynamic variables (such as laboratory test results and vital signs) contribute significantly to the 8-hour horizon predictions, they are not as important to the 24-hour predictions. Static variables (either demographic variables or measurements from the time of admission) also contribute significantly to both models. The detailed Shapley Additive Explanation plots for each task are provided in Figure 3 and the exact definition of variables included in each class of features are described in Section 2 of the Supplementary Information.

**Figure 4:**
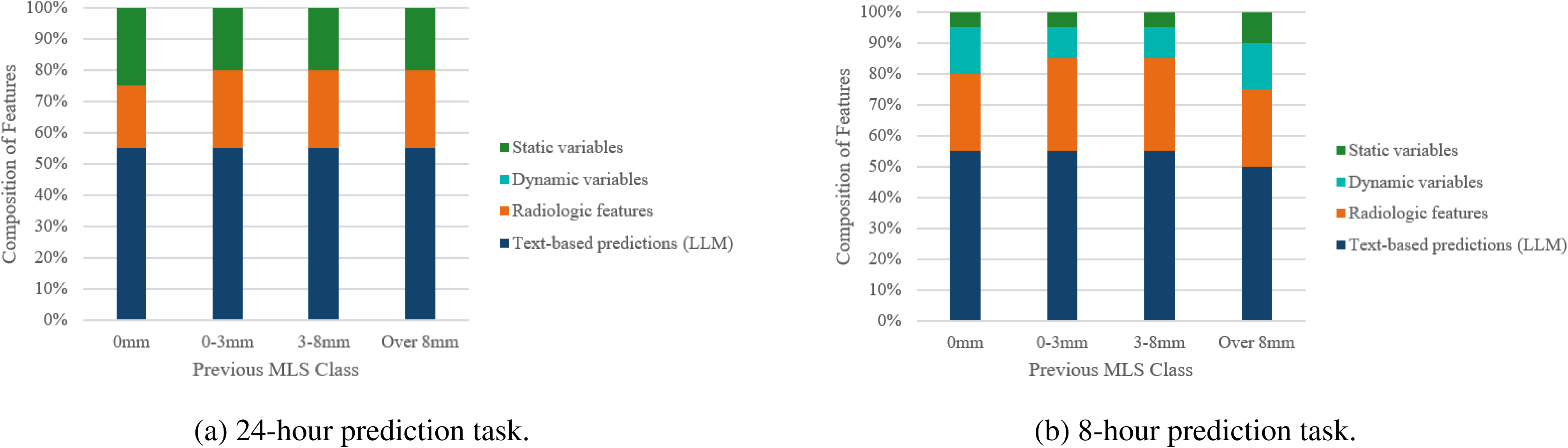
Comparison of feature composition by previous MLS class for the downstream models trained on 24-hour and 8-hour prediction tasks.

In the 24-hour prediction task, text-based predictions generated by the large language models emerge as key determinants of patient trajectory. Expert-curated radiographic measurements, including the most recent MLS value, the prior MLS value, and the time and value of the first measured MLS, also contribute significantly to HELMET predictions. Our analysis also highlights the role of approximate stroke size based on NIHSS, as well as laboratory markers, such as maximum white blood cell count over the past 24 hours and blood urea nitrogen at admission. Notably, the administration of 23.4% hypertonic saline is the only treatment-related feature ranked among the top predictors. Furthermore, patient age becomes a significant factor in predicting edema progression within higher MLS classes.

In the 8-hour prediction task, the most recent and prior MLS values, along with class probabilities derived from the large language model, are the most significant contributors. Similar to the 24-hour model, the timing and value of the first MLS measurement, as well as the time of the first MLS value exceeding 3 mm, rank among the top features. Laboratory markers, including white blood cell count and blood glucose levels, alongside vital signs such as pulse and temperature, further aid in distinguishing between edema states. Notably, no treatment indicators are among the highest-ranked predictors.

## 3 Discussion

Our dynamic time series models leverage non-linear machine learning algorithms to predict the risk of edema and the trajectory of MLS using a comprehensive multi-modal dataset for two distinct future time windows. To the best of our knowledge, HELMET-8 and HELMET-24 constitute the first dynamic risk models for predicting the trajectory of cerebral edema on an hourly basis, leveraging data from two healthcare systems.

Existing cerebral edema risk scores provide only static predictions of late clinical outcomes, such as death or decompressive hemicraniectomy, primarily using linear techniques [17–20] (with some use of non-linear models [29]). While useful for initial triage, these models are limited in their utility as clinicians follow individual patients over time and make decisions based on new data. Recent work by our group has highlighted the significance of incorporating post-baseline patient data to enhance the prediction of inpatient outcomes [21]. No studies to our knowledge had utilized granular updating information over the course of hospitalization to estimate the actual state of cerebral edema by objective measurements, such as MLS.

Our analyses in the derivation cohort reveal that HELMET-24 outperformed HELMET-8 by a small margin across all overall metrics. However, the performance difference across tasks was more pronounced when looking at filtered metrics, indicating that the HELMET architecture may be better suited for making predictions over longer time windows. One possible explanation for this observation is that changes in edema trajectory can appear to occur more abruptly when looking at closer time horizons, while such changes in edema trajectory are smoothed over longer prediction windows. Another key contributing factor is likely the frequency of imaging, the primary input feature of our models, which was obtained approximately every 11.3 hours in the derivation cohort and every 18.8 hours in the validation cohort. This difference in scanning frequency between the cohorts likely also explains the improved performance of HELMET-24 over HELMET-8 in the external validation cohort. Information on the frequency of laboratory and vital sign data collection is reported in Section 2.2 of the Supplementary Information. Subsequent studies could elucidate optimal predictive horizons for these newly developed hybrid models.

By incorporating predictions from fine-tuned large language models into HELMET, we advance the existing literature on multimodal machine learning in medicine [30, 31]. Our models were significantly improved by the inclusion of both upstream predictions from large language models fine-tuned on raw radiology reports as well as manually-measured neuroanatomic variables hypothesized as relevant by neurology experts (see Figure 2). Our hybrid approach highlights the benefit of using clinician-generated raw texts to capture otherwise unmeasured variables and physician beliefs about the patient’s trajectory. However, while features derived from the large language model predictions make up a plurality of high-impact features across both tasks (see Figure 4), the large language model predictions in isolation were inadequate indicators of MLS trajectory (see Section 2.4 of Supplementary Information). Our findings underscore that multi-modal hybrid approaches combining both expert-derived features and raw data appear to significantly enhance outcome prediction accuracy in clinical settings and may inform which features should be routinely included in radiology reports.

The Shapley Additive Explanation analysis (Supplementary Information Figure 3) enables us to infer key clinical variables that drive model performance. Dynamic variables capturing previous MLS measurements were the most critical predictors of future MLS status. Additionally, key time-related features, such as the time of the first non-zero MLS and the first MLS exceeding 3 mm, highlight the natural course of edema growth, which typically peaks between 2-5 days post-ictus [7, 11, 13, 32–34]. Time is a crucial factor often overlooked in other studies. While clinicians often assess risk based on time subjectively, our model is the first to quantitatively integrate time in a standardized way. In contrast to most risk algorithms that neglect temporal dynamics, our results demonstrate that accurate time quantification is essential for predicting imminent edema progression, aligning with clinical intuition that the same degree of edema is less critical later in the treatment course. In the absence of clear guidelines on imaging timing and its influence on surgical outcomes [9], our model provides valuable guidance for clinicians in predicting MLS worsening and optimizing patient management using real-time data.

Consistent with the literature, our Shapley Additive Explanation analysis highlighted other influential variables, including dynamic white blood cell count, admission blood glucose, and temperature. Elevated glucose at presentation has been previously linked to malignant edema and poor outcomes [17], while recent analyses showed that white blood cell count and temperature increase before radiographic evidence of mass effect [26]. Our results also highlighted that laboratory test results and vital signs have higher predictive power in the 8-hour compared to the 24-hour task, capturing more effectively short-term changes closer in time to the event. Our finding that hypertonic saline administration was also among the top features for HELMET-24 likely reflects the subjective physician risk assessment of the patient and, due to the observational nature of the data, does not provide any causal insights. Intriguingly, we did not observe similar importance of mannitol or other preparations of hypertonic saline, which may reflect its use in clinical practice and should be further studied. Given the heterogeneity in medical treatment patterns [9], the connection between osmotic therapy and clinical intuition should be interpreted with caution.

Our results reveal that human insights and radiographic features extracted from scan images play a complementary role to the large language model predictions, despite using the same foundational data source (CT images). Medical professionals bring nuanced understanding through expert-curated radiographic features, adding a layer of interpretability and context that purely algorithmic approaches may lack, especially in settings with limited sample size. Complementing past studies focused on human-AI interactions [35, 36], our work underscores the significant role of human insights in enhancing the predictive power of large language models, particularly in scenarios where critical variables, such as MLS, are not routinely recorded in structured form. The importance of such specific expert-curated radiographic features in our models indicates a possible improvement to radiology reporting whereby clinicians should aim to extract and record additional radiographic features at the time of observation in their report texts.

The synergy between algorithm-derived insights from large language models and human-extracted features highlights the potential of hybrid artificial intelligence systems in clinical settings. These systems leverage the precision and scalability of machine learning while retaining the critical contextual understanding provided by human expertise. This combination is especially valuable in settings such as neurocritical care, where the dynamic nature of conditions like cerebral edema demands a nuanced approach to prediction and intervention. In such highly specific tasks, and in the absence of large databases for research, the successful creation of robust machine learning models may hinge on the consistent and coherent extraction of features. Researchers should aim to work with clinicians to generate more robust datasets of human-extracted variables needed to develop better prediction tools, and future studies in clinical machine learning can benefit from using such datasets by employing hybrid approaches.

Notably, the HELMET models generalize to an external population substantially better than previous baseline models [17, 18]. While there is an expected decrease in performance from all internal validation to external cohorts due to unmeasured, context-dependent parameters, our results demonstrate that model generalizability can be improved by leveraging dynamic features. When deployed to a new site, we anticipate that HELMET, like other models, will benefit from re-calibration for maximally effective use. Nevertheless, our external validation results enhance confidence in the applicability of HELMET, as well as the broader use of our dynamic, hybrid approach for developing risk prediction models in clinical practice.

An important implication of our work is that shifting risk prediction from a single baseline assessment to a continuous, longitudinal approach could improve real-time patient triage, optimize imaging resource use, and explore whether early-warning alerts based on these predictions can enhance patient outcomes and care quality. Studying the implementation of these algorithms will be crucial in assessing their clinical utility. Future research could build on existing studies showing the effectiveness of machine learning tools when paired with well-designed clinical interventions. Additionally, sensitive models like these could have a significant impact in smaller or under-resourced settings where neurointensive care may be unavailable or overburdened. Further investigation is also needed to bridge the gap between prediction and action by using machine learning-derived predictions to provide prescriptive recommendations for when scans or clinical interventions should occur.

### 3.1 Limitations

There are several limitations to our work. The dataset size was constrained by the specific inclusion criteria and the manual labeling of radiographic images. Since imaging was performed at the discretion of treating physicians, the timing of MLS measurements was inconsistent. While MLS is just one indicator of worsening mass effect, and its clinical relevance may vary depending on factors such as age and brain atrophy, it remains a well-established, critical, and measurable radiographic biomarker of cerebral edema.

Similar to other retrospective studies, our dataset contains missing values due to its reliance on retrospectively collected hospital electronic health records, which were imputed using widely established unsupervised learning techniques. We lacked access to key physical examination data, such as neurological deterioration, quantitative pupillometry, and other longitudinal multimodal monitoring methods including electroencephalograms, optic nerve sheath diameter, or direct intracranial pressure monitoring. Further, we lacked functional outcome data from the end of a patient’s hospitalization, including modified Rankin score values. The integration of these variables is a clear focus for future direction. However, these tools are not ubiquitous among all hospital centers or are not routinely used in clinical care (such as intracranial pressure monitoring in ischemic stroke), and therefore, using variables available to most centers leads to broader model generalizability and utility. We also acknowledge that there was a significant increase in reported observations per patient after 2016 due to a change in the electronic medical record system. This created two quasi-distinct data distributions within the training set. We believe that the successful generalization to external validation cohort data suggests that both data distributions are valuable.

Temporal transformations in the data may introduce bias, as patients with more complete data have multiple observations. Our approach, which uses the most recent and maximum values while compact, may oversimplify the trajectory of MLS measurements. Even though our hospital systems differ significantly in racial and socioeconomic composition, inter-regional generalization remains uncertain. Finally, despite the multi-year gap between data collection in the derivation and external validation cohorts, this study remains retrospective. Further prospective clinical studies are needed to validate real-time predictions and assess their impact on clinical practice.

## 4 Methods

Figure 2 illustrates the study design, including dataset curation, feature extraction, and model development. The summary diagram presents how the multimodal datasets were constructed, transformed, and used for model development and evaluation. Section 4.1 outlines the data sources and patient selection criteria (Panel A). Section 4.2 describes the dataset curation and feature transformation process (Panels B and C), and Section 4.3 details the model training and evaluation process for the HELMET and EDEMA baseline models (panel D).

### 4.1 Patient Identification & Exclusion

The derivation cohort originated from the Massachusetts General Brigham hospital system research patient data registry and electronic data warehouse, which form the centralized clinical data registries of the organization. The external validation cohort was obtained from the Boston Medical Center electronic medical records (see panel A of Figure 2). The patient inclusion diagrams for both cohorts are shown in Figure 1.

For the retrospectively identified derivation cohort, we queried the Massachusetts General Brigham data registry for unique patient encounters between January 8^th^, 2006 and July 5^th^, 2021 with stroke diagnosis codes and used an established natural language processing model to identify patients with acute MCA stroke involving *≥*1/2 of the MCA territory [37, 38]. For the prospectively collected external validation cohort, we leveraged an existing registry of patients with acute MCA ischemic stroke involving at least 1/2 of the MCA territory admitted after May 15^th^, 2019 and discharged before November 25^th^, 2023 who also had pupillometry measurements. We excluded patients with confounding injuries^3^, including acute contralateral or posterior fossa injury; encephalomalacia exceeding 1/2 of the vascular territory; extraparenchymal, subarachnoid, epidural, or subdural hemorrhage; moyamoya disease; and cerebral venous sinus thrombosis. Patients were also excluded for having an unknown last-seen-well date or a last-seen-well date more than 24 hours before presentation. In the external validation cohort, we also excluded patients for whom life-sustaining treatment was withdrawn within 24 hours of admission without an interval CT scan and those whose pupillometry data did not meet established criteria^4^. Finally, patients from either cohort were excluded for not having radiographic imaging data after the initial 24 hours of hospitalization.

### 4.2 Data Curation

We used demographic, clinical, and text-based variables at varying time horizons to construct the HELMET models. These variables include time-invariant demographic variables, clinical variables recorded at the time of admission, time-censored dynamic clinical variables changing throughout admission, and radiology reports generated by clinicians at the time of each true scan. The multi-modal structure of the datasets and the operations applied to create the final features are illustrated in panel B of Figure 2. Further detail regarding the data curation, feature extraction, and missing data imputation process is available in Section 2 of the Supplementary Information.

#### 4.2.1 Variable Processing

Demographic variables included age and sex. Race and ethnicity were not used as inputs to the HELMET models in order to reduce the risk of introducing discriminatory bias into the predictions. Other static admission-related variables included the time of admission, time last seen well, past medical history (including prior stroke, hypertension, and anticoagulant or antiplatelet use), vitals and laboratory blood value readings taken on admission, and the NIHSS score. Static variables from patient medical records accounted for 33 features in the final datasets. Dynamic clinical variables included 16 features, including vital sign data, laboratory test results, and treatments administered. Features were selected from the patient medical record due to their possible association with malignant edema [1, 8, 39, 40]. Radiographic variables were collected by neurology-specialized team members who labeled the radiographic images at the time of dataset construction. The main variables of interest from the radiographic images were the size of MLS and the size of the pineal gland shift from each scan. We also extracted the time and value of measurements exceeding certain clinically relevant thresholds, such as the first MLS value over 3mm. A total of 36 human-labeled radiographic variables were included in the dataset (see for further information Section 2.3 in the Supplementary Information).

#### 4.2.2 Data Restructuring

Feature data was aggregated into hourly intervals, and missing values for static variables were imputed using the mean of each feature column, employing the SciKit Learn Simple Imputer [41]. To increase the number of trainable observations per patient, multiple hourly observations were generated, starting from the time of admission and continuing until either discharge or surgical intervention (decompressive hemicraniectomy). Each observation hour was re-indexed based on the number of hours since the patient was last seen well. Static variables were carried forward across all hourly observations, while dynamic variables were assigned to observations corresponding to the specific hour at which they were recorded.

Given that each observation was constructed on a single-hour interval, data from previous hours was absent in subsequent observations, and any variables not measured within a particular hour were treated as missing. To enhance model performance, we incorporated historical information into each observation by applying time-based transformations of the dynamic variables. Specifically, time varying features from both electronic medical records and radiographic imaging were carried forward using two methods, resulting in two derived features per original variable: one representing the maximum value over the previous 24 hours (rolling window), and the other capturing the most recent available measurement (forward-fill). Panel C of Figure 2 provides an illustration of this process. These transformations yielded 32 features from 16 original medical record variables and 72 features from 36 original radiographic variables. Additionally, a 73^rd^ forward-filled feature was introduced to capture the last known MLS value prior to the most recent measurement, enabling better characterization of the patient’s trajectory. The full lists of variables used in training HELMET-8 and HELMET-24 with feature definitions can be found in Tables 5 and 6 in the Supplementary Information.

#### 4.2.3 Target Outcome Construction

We chose future MLS as the outcome of interest due to its critical role in determining the severity of ischemic stroke [17, 20, 42]. We divided the continuous range of MLS values into discrete MLS categories, chosen based on input from collaborating physicians as to the most clinically useful thresholds corresponding roughly to no, mild, moderate, and severe MLS. While 5mm of MLS has been previously used in early models to define severe edema [1], our dataset revealed that MLS typically approached 8mm before decompressive hemicraniectomy, leading to the choice of 8mm as the upper threshold of the prediction classes. Other research has shown >3mm may be associated with worse outcomes [12]. Based on this evidence, 3mm was set as the middle cutoff of our edema classes. We therefore employed the MLS categories (classes) of no MLS [0mm], MLS of less than 3mm (0-3mm], MLS between 3mm and 8mm (3-8mm], and MLS exceeding 8mm (>8mm).

We defined two prediction targets: the maximum MLS value within the subsequent 8-hour window and the maximum MLS value within the subsequent 24-hour window. These intervals were selected for their clinical relevance, providing a balance between the need for timely diagnostic and therapeutic interventions and the typical cadence of updated clinical data, including laboratory results, imaging studies, and vital signs. The target MLS values were derived as the maximum MLS recorded from radiographic images within the specified prediction window and subsequently mapped to one of four pre-defined MLS classes, thus creating two distinct classification tasks for each observation.

To ensure target validity, we excluded from our analysis observations for which no radiographic scan occurred within the relevant prediction window (e.g., for a 24-hour prediction task, if there was a 40-hour gap between scans, the first 16 hours post-scan were excluded as valid targets could not be constructed). This approach resulted in the generation of up to eight observations per scan for the 8-hour prediction task and up to 24 observations per scan for the 24-hour task.

#### 4.2.4 Large Language Model-Derived Features

Radiology reports, written by radiologists at the time of hospitalization as interpretations of CT scans, offer clinical insights not captured by the quantitative measurements of MLS and pineal gland shift alone. The Clinical-Longformer [43], initially trained on large corpora of clinical text for general medical language modeling, was adapted for multi-class classification using the radiology reports and their corresponding future MLS class labels. This particular pre-trained transformer model was selected based on a review of recent literature [44]. By specifying the intended task of text classification when loading the pre-trained model, a linear classification layer was added at the model head to transform the default output into predictions on the four-class MLS ranges. During fine-tuning, the model’s pre-trained weights were updated by minimizing the categorical cross-entropy loss between the predicted class probabilities and the true MLS class labels over 8-hour, 24-hour, and 36-hour^5^ prediction windows. Although the clinically observed time intervals of observation are shorter, the 36-hour targets are used only in the context of feature extraction from the radiology reports to capture longer-term MLS trajectories. By leveraging the pre-trained language model’s understanding of clinical terminology and combining it with task-specific data, we enabled the model to effectively learn nuanced patterns in the radiology reports relevant to future MLS prediction. The per-class probability outputs from each of the three large language model classifiers were then added as features to the dataset used in training the HELMET models, resulting in 12 additional features (one for each of the four classes from each of the three models). Further details of the large language model fine-tuning and performance are provided in Section 2.4 of the Supplementary Information.

### 4.3 Predictive Model Development

To derive HELMET-8 and HELMET-24, multi-class classification models were trained to predict the 8-hour and 24-hour MLS trajectories. By discretizing continuous MLS values into four predefined classes, the models reframed the regression task into a classification problem, predicting into which of the four MLS ranges a patient’s maximum MLS would fall during the specified prediction window.

#### 4.3.1 HELMET Model Derivation

The HELMET models are trained using the XGBoost algorithm [45], a well-established ensemble learning technique suitable for multi-class classification that leverages tree-based gradient-boosting. We also explored the use of alternative methods, but we opted to use XGBoost as we did not see any significant changes in downstream performance (see Section 4.2 in the Supplementary Information). We divided the derivation data using five-fold randomized splits. Data were partitioned at the patient level to ensure there was no leakage of observations from the same patient in the training and testing sets of the derivation cohort. Each HELMET model was trained on data from 80% of the patients in the derivation cohort in each of the five splits. Models were then evaluated on data from the remaining 20% of the Massachusetts General Brigham patients for internal validation, and all Boston Medical Center data for external validation. We aggregated performance metrics across all randomized data partitions, allowing us to report confidence intervals around each averaged performance metric.

To prioritize accurate identification of MLS worsening, the algorithm training was based on a modified cross-entropy loss function that gives higher importance to observations with target values different from the immediately preceding MLS class.^6^ We refer to these transitioning observations as the “filtered” dataset, with the set of all observations (transitioning and non-transitioning) referred to as the “overall” dataset. The non-transition weight value was tuned as a model hyperparameter.

Leveraging the Weights&Biases machine learning training platform [46], we applied Bayesian optimization to fine-tune the hyperparameters of the HELMET models, utilizing filtered AUROC as the optimization objective function [47]. The resulting values of the HELMET models hyperparameters are detailed in Section 4.3 of the Supplementary Information. We also conducted several sensitivity analyses to test our model structure and development methods, which are explained in further detail in Section 4.4 of the Supplementary Information. All computational experiments, including model development, validation, and evaluation, were performed using Python 3.11 and the Scikit Learn library [41].

#### 4.3.2 EDEMA Baseline Derivation

To compare HELMET with a baseline, we opted to use linear models akin to the models commonly reported in the existing literature [18, 19, 42]. Specifically, we developed multinomial regression models that leverage similar independent variables to the EDEMA score [17]. The EDEMA score is a multinomial regression model developed for predicting the adverse event of malignant edema after stroke, leveraging the following variables as input: basal cistern effacement, admission MLS, glucose, previous stroke, and the use of medical thrombolysis or thrombectomy interventions. We approximated the EDEMA score by training multinomial regression models in the derivation and external validation cohorts to predict our target outcomes of interest. By separately training the baseline models to each cohort and prediction task, we derive a different model for each dataset leading to four total baseline models: Massachusetts General Brigham EDEMA-8 and EDEMA-24, as well as Boston Medical Center EDEMA-8 and EDEMA-24 (see Figure 2). Our baseline models use as input the static features of blood glucose at admission, HbA1C at admission, history of previous stroke, mechanical thrombectomy at any point, and medical thrombolysis using tPA at any point, as well as the dynamic features of most recent blood glucose, presence of basal cistern effacement on the most recent scan, and the MLS measurement from the most recent scan (indicated by asterisks in Table 1). The baseline models were derived using five-fold splits at the patient level for both the derivation cohort and the external validation cohort. For training the EDEMA-8 and EDEMA-24 for Massachusetts General Brigham and the HELMET models, we used the same train-test partitions to maximize comparability.

### 4.4 Model Evaluation

#### 4.4.1 Performance Evaluation

The principle target metric of model evaluation during algorithm tuning was filtered AUROC. We selected this criterion given its relatively universal use as a performance indicator [48, 49] and its suitability for use in comparison between different model types. To evaluate the predictive performance of the derived models, we report the downstream AUROC, AUPRC, accuracy, sensitivity, and specificity of the overall and filtered datasets across both the testing set of the derivation cohort and the external validation dataset. We separately evaluated model performance on the filtered cohort to stress the clinical importance of cases where a patient’s MLS class changed. All performance metrics are reported as the mean and the corresponding 95% confidence intervals for the testing sets of the derivation cohort and the external validation cohort.

AUROC and AUPRC are typically defined only for binary classification models. Given that our targets belong to a four-category classification task, we calculated the true positive rate, false positive rate, sensitivity (recall), and precision scores in a one-versus-rest binary classification setup across prediction thresholds ranging between zero and one [41, 50]. The resulting metrics were then averaged across the four classes at each given threshold to derive the receiver-operator characteristics and precision-recall curves weighted by the number of observations in each class. Accuracy was defined as the proportion of observations where the model correctly assigned the highest predicted probability to the true target class. To adapt sensitivity and specificity for the multi-class setting, we reformulated the task to focus on correctly predicting whether a patient’s MLS state would worsen (i.e., increase) within the prediction window. For this binary classification task, true labels were assigned a value of one if the future MLS class exceeded the current MLS class, and zero otherwise. Predicted values were similarly assigned a value of one if the MLS class with the highest predicted probability was greater than the current class, and zero if it was not. Sensitivity and specificity were then computed according to their standard definitions.

#### 4.4.2 Feature Importance Analysis

We utilized Shapley Additive Explanation analysis to study the relative importance of various features in our final ensemble learning models [51]. The Shapley values measure the marginal contribution of each feature to a prediction in a machine learning model by decomposing the prediction into the sum of effects from each feature, utilizing the principles of cooperative game theory. For each class prediction task, Shapley values were computed to identify the features most influential in predicting across the four MLS classes. Per-class Shapley values were aggregated to generate a ranked list of overall feature importance. While the absolute Shapley values are not directly informative, the relative ranking and composition of top features offer valuable insights into model behavior and potential implications for future clinical practice and edema prediction.

### End Material

#### Ethics & Inclusion

Ethics approval was obtained from the Institutional Review Boards of Massachusetts General Brigham and Boston Medical Center. Informed consent was not required because the only patient data collected was standard of care, no research intervention was implemented, and the study used anonymized, retrospective patient records.

To promote the inclusion of diverse and historically marginalized populations, we sourced external validation data from a more racially and socioeconomically diverse patient population. Additionally, race and ethnicity were not used as inputs to the HELMET models in order to reduce the chance of introducing racial bias into the models.

#### Guidelines

Transparent Reporting of Multivariable Prediction Models for Individual Prognosis or Diagnosis and Journal of Medical Internet Research Guidelines for Developing and Reporting Machine Learning Predictive Models in Biomedical Research were followed.

## Code Availability

The code developed for this study will become publicly available upon publication of the manuscript by a peer-reviewed journal. All model development and testing was done using Python 3.11 and publicly available packages (sklearn, pandas, numpy, wandb, xgboost, datasets, transformers, evaluate, torch, cupy, fire).

## Author Attribution

EP co-led model development and evaluation, led data analysis, and drafted and edited the manuscript.

OOD co-led initial model development and evaluation and assisted with the manuscript revision.

YZ organized the results and figures and revised the manuscript.

PT proposed ideas and worked in the theoretical underpinnings of this paper.

LAM participated in data collection and curation and assisted with manuscript revision.

JP participated in data collection, dataset creation and adjudication.

SC participated in data collection, adjudication, dataset creation and manuscript revision.

YD participated in data collection, dataset creation and manuscript revision.

JS participated in data collection, dataset creation, and adjudication.

BB participated in dataset creation, data adjudication and manuscript revision.

SS co-led study design and scope; provided clinical insight; and participated in manuscript review.

CJO co-led study design, data collection, data curation, and interpretation and provided clinical insight. CJO contributed to interpreting the findings and drawing conclusions about the model’s effectiveness. CJO actively participated in drafting and review.

AO co-led the study design and coordination and guided the data preprocessing and the machine learning model development and training. AO contributed to interpreting the findings and drawing conclusions about the model’s effectiveness. AO actively participated and led the manuscript writing and revision process.

## Data Availability

The data used in our study come from two academic medical centers in the United States subject to the Health Insurance Portability and Accountability Act. Due to the data use agreement that we have signed with Boston Medical Center and Massachusetts General Brigham, the datasets cannot be made publicly accessible, as they contain protected health information and other sensitive information about the patients. Any user that wishes to gain access to the dataset needs to become HIPAA certified and get approved as an authorized by the collaborating healthcare systems Institutional Review Boards.

---

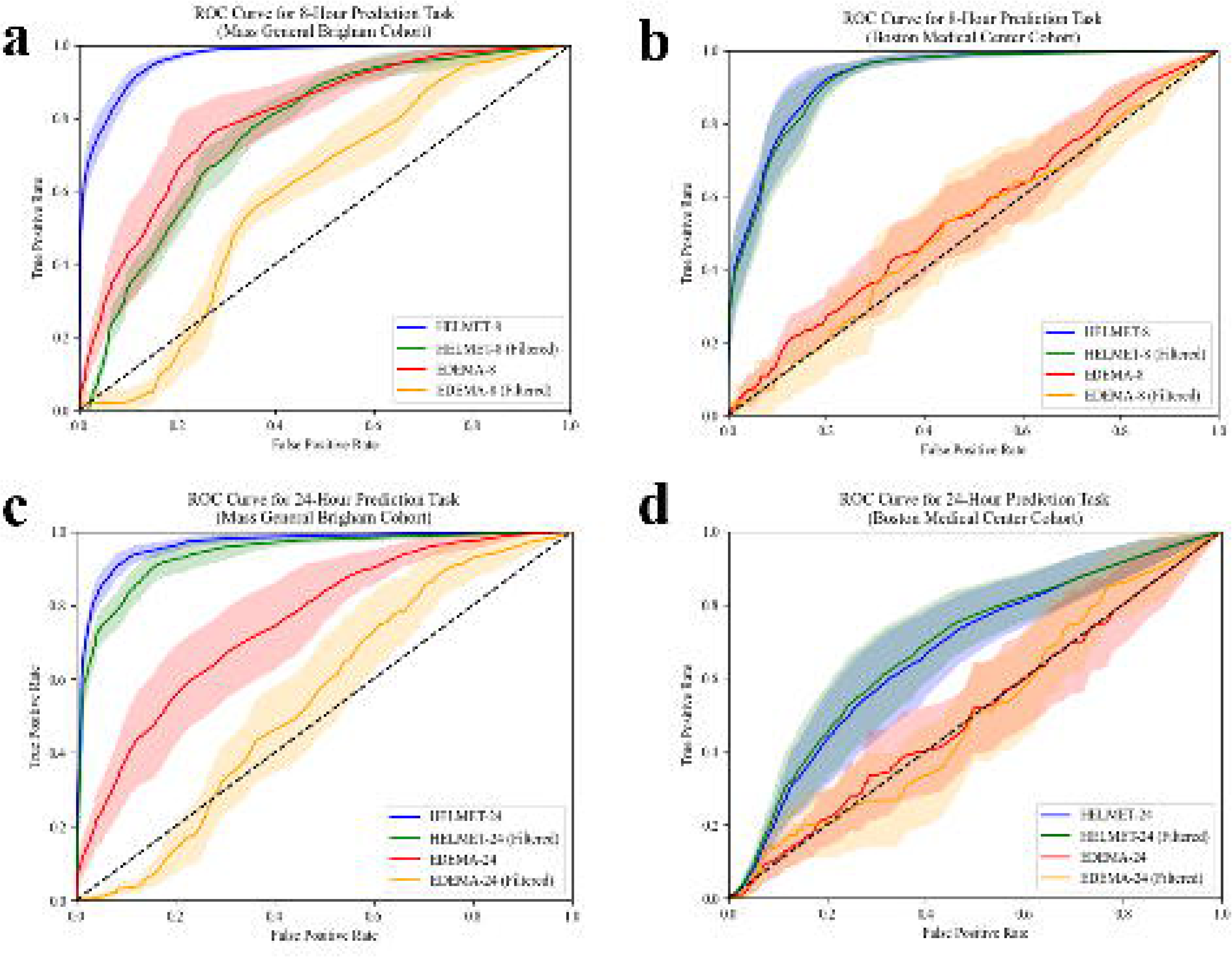

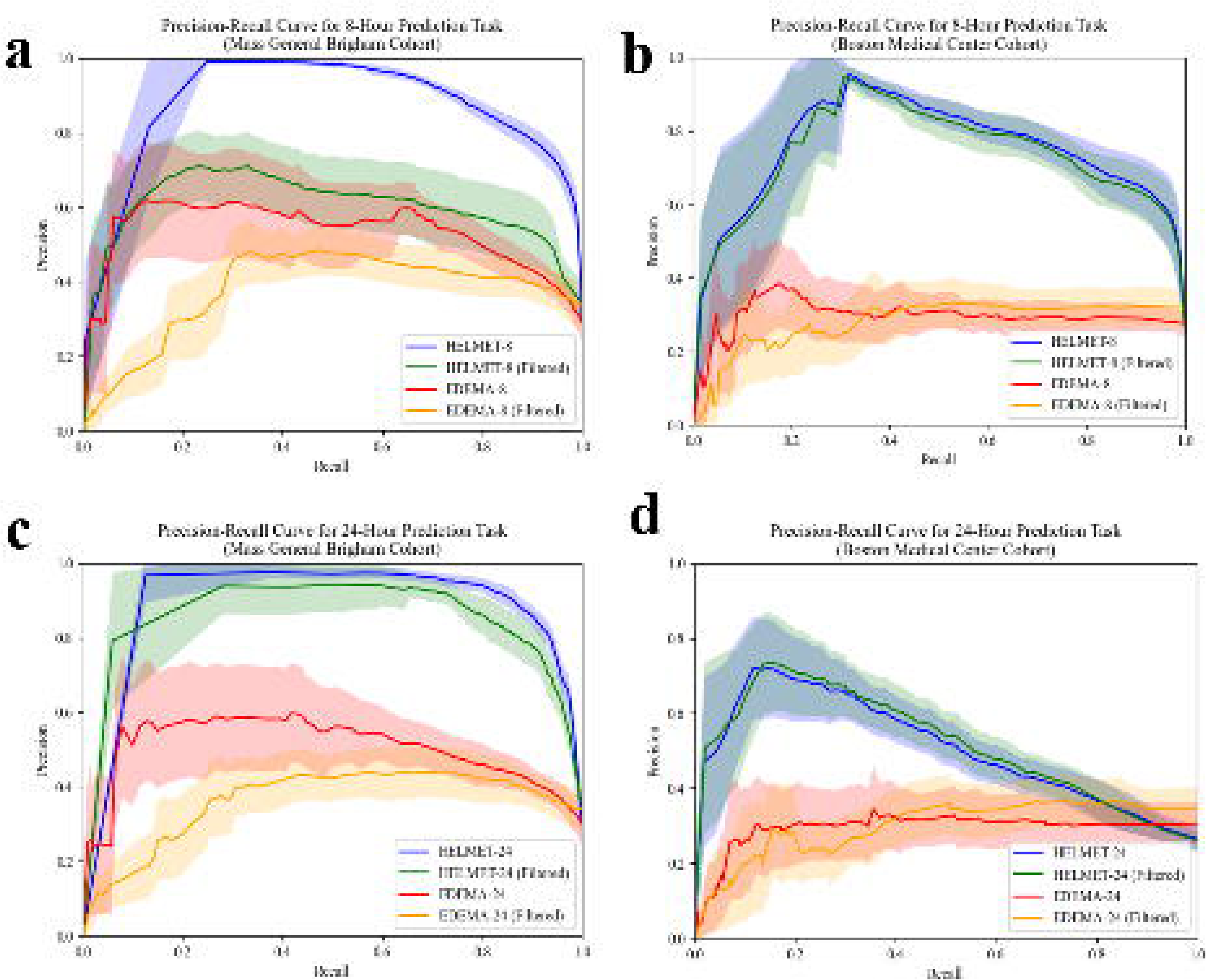

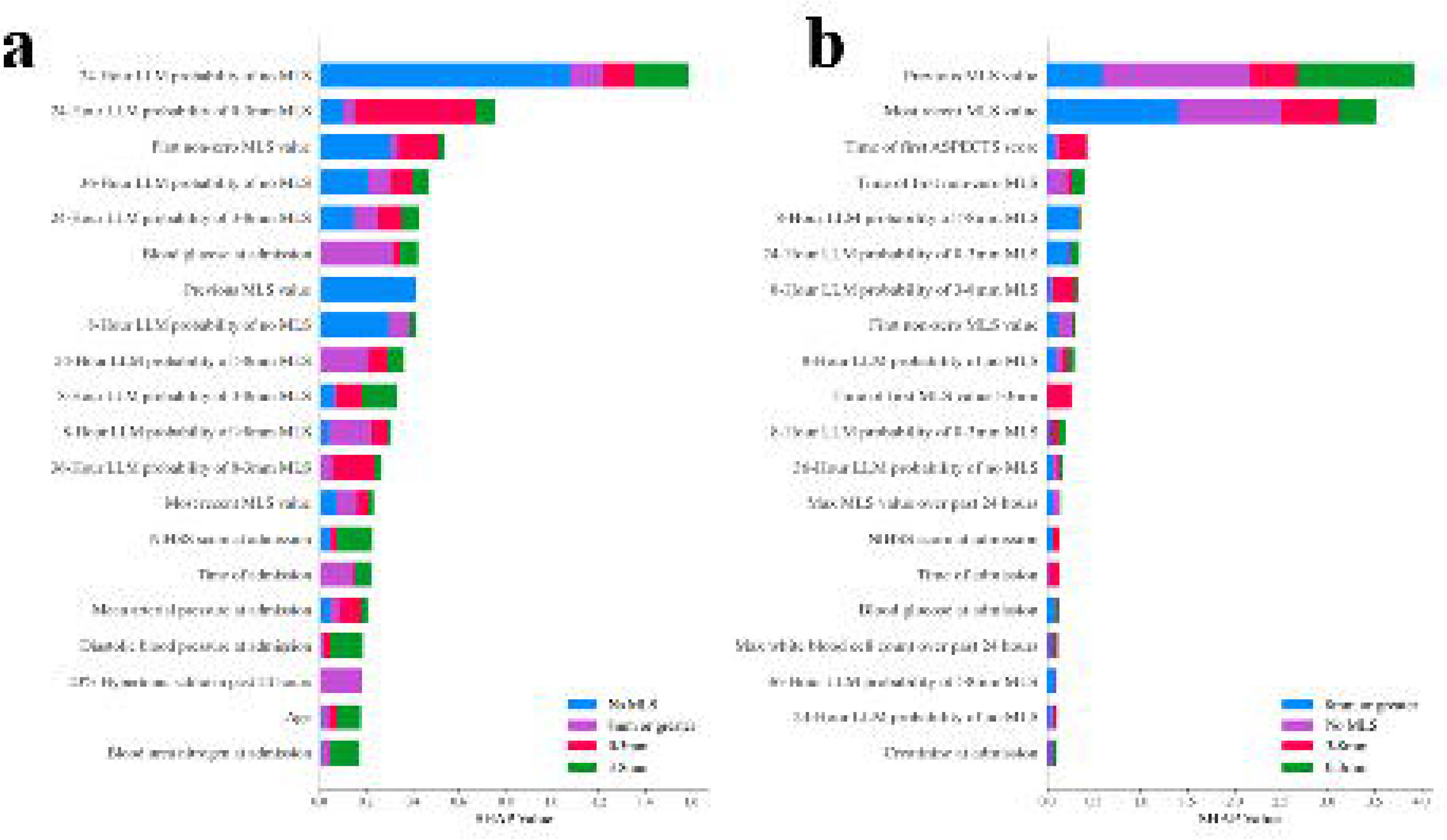

## Footnotes

1 By design, the EDEMA baseline models were provided an edge over the HELMET models on the external validation cohort since they were trained separately on both the derivation and external validation datasets, while the HELMET models were only trained on the derivation dataset.

2 Since our classification problem required prediction of future MLS among four possible classes, a random guess model would result in an average AUROC of 0.5, average AUPRC of 0.25, accuracy of 0.25, sensitivity of 0.25, and specificity of 0.75. Therefore, performance scores exceeding these thresholds are described as better than random. The exact formulation of composite average metrics is described in Section 4.4.1.

3 For patients who had hemicraniectomies, data were omitted after a hemicraniectomy had occurred.

4 Established pupillometry data criteria included having a minimum of three observations prior to MLS ≥5mm or other evidence of mass effect, no data gaps exceeding 24 hours, no history of conditions which might effect pupillometry data, and initially reactive and normal pupils.

5 36-hour training targets corresponding to the maximum MLS reached over the following 36-hour were derived using the same process described above for the 8-hour and 24-hour targets but were only used in training the large language model classifiers.

6 The modified cross-entropy loss function incorporates both filtered and non-filtered observations to account for transitions in the patient’s state. Let *N* represent the total number of observations, and *C* be the total number of classes. For each observation *i* ∈ {1, *…, N* }, *yi* ∈ {0, 1}*^C^* is the one-hot encoded true label vector, where *yi,c* = 1 if observation *i* belongs to class *c*, and *pi,c* denotes the predicted probability that observation *i* belongs to class *c*. Let F ⊆ {1, *…, N* } represent the set of filtered observations, where a patient’s state changes over time, and N F = {1, *…, N* } \ F denote the non-filtered observations, where no state change occurs. The loss function is given by 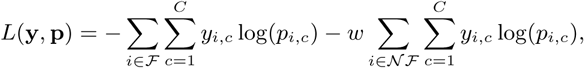 where *w* ∈ (0, 1] is a weight applied to reduce the contribution of non-filtered observations. The first term represents the cross-entropy loss for filtered observations, and the second term represents the weighted cross-entropy loss for non-filtered observations. The final values of the weighting term, *w*, for both models can be found in Table 8 in the Supplementary Information.

