## Supplementary Information for "HELMET: A Hybrid Machine Learning Framework for Real-Time Prediction of Edema Trajectory in Large Middle Cerebral Artery Stroke"

### 1 Review of Existing Prediction Models

We present five existing models for predicting malignant MCA infarction or malignant brain edema. All of the reviewed models use logistic regression to make static predictions of late-stage patient outcomes, and are therefore limited in their applicability to real-time medical decision-making. The five existing models are summarized in Table 1.

### 2 Data Collection and Processing

Data from Mass General Brigham were extracted from the hospital's electronic medical records database using the Research Patient Data Registry and the Electronic Data Warehouse. This included demographic and outcomes for structured data, as well as radiology reports and clinical notes for unstructured data. A second researcher verified all data to ensure accuracy. Similarly, data from Boston Medical Center was obtained using the hospital's electronic medical records database using Clinical Data Warehouse which included structured and unstructured data as well. Decision rules regarding ascertainment and cleaning of all data from the electronic health records are described in Pohlman et al, 2024 [1].

#### 2.1 Stroke Characteristics

Last seen well date and times were identified via clinical notes. Last seen well was defined as the last time before a patient experienced a substantial deficit as indicated by the NIHSS (symptoms of numbness, tingling, or other symptoms that would result in NIHSS < 3 were not taken as the time of last seen well). Instances where last seen well lacked exact times or only referenced time were labeled as "inferred." If last-seen well occurred prior to an in-hospital surgery, the surgery time was identified. If no surgery start time was identified, surgery was assumed to be at 7:00AM. If last seen well referred to "last night before bed," with no exact time, then last seen well was assumed to be 10:00 PM. All last-seen well data underwent supervisor review (CJO).

*NIHSS* was extracted from history and/or physical exam. In cases where multiple NIHSS were reported, the last one (after any procedures took place) was used. NIHSS scores were documented via clinical notes. For patients who underwent medical or mechanical thrombolytic procedures, we prioritized the NIHSS recorded after intervention.

*Vessel Occlusion location:* All CT Angiograms were reviewed and classified as a vessel occlusion based on radiology reports. The most proximal vessel involved (Internal Carotid Artery, M1, M2, M3/M4) was used for the vessel occlusion location. The anterior cerebral artery was also identified as occluded or not. In instances in which reports described vessel occlusion but did not explicitly name vessel occluded, M1 was assigned if occlusion was described to begin prior to bifurcation/trifurcation.

#### 2.2 Laboratory and Vital Sign Measurements

Laboratory and vital sign values were obtained through the structured Research Patient Data Registry/Clinical Data Warehouse database. The first value present within eight hours before and up to thirty-six hours after presentation date time was used, accounting for labs that occurred prior to the first radiographic scan and/or clinically determined presentation date time. The mean time from presentation to first laboratory value in the derivation cohort was 0.31 hours (approximately 19 minutes), with a standard deviation of 17.9 hours. Laboratory values taken prior to presentation time are due to presentation time definition—labs in the emergency department occasionally predated official "presentation time." Vital signs at presentation were derived from a chart review of the History and Physical. On average, vital signs data were updated every 51.0 hours (standard deviation 38.9 hours) and laboratory data were updated every 64.1 hours (standard deviation 40.3 hours) within the first week after presentation.

To address clearly erroneous lab or vital sign errors, each longitudinal measurement was assigned a Z-score based on the data distribution across the entire cohort. Measurements with an absolute Z-score of three or greater, or those showing a 25% change from a preceding value, were flagged as a potentially erroneous measurement. The principal investigator reviewed each patient's flagged laboratory/vital sign trajectories within the context of their hospital course to determine which outliers were valid and which needed to be removed. Temperature values recorded in Celsius (30°s-low 40°s) were converted to Fahrenheit. Other abnormally low-temperature values, such as 78°F, were removed. One sodium value of 102 mEq/L was removed after chart verification. All glucose values $\geq$1000 mg/dL and one glucose measurement of 866 mg/dL were removed after chart verification.

#### 2.3 Radiographic Features

The population's radiographic features were derived using a Natural Language Processing algorithm that screens for patients with acute MCA stroke identified via ICD-9, ICD-10 codes [2]. Patients with large acute MCA stroke were confirmed to be $\geq$ 1/2 of the MCA territory by designated, trained M.D members of the research team (SC, CJO). Images included in $\geq$1/2 MCA territory had to have either full superior or inferior MCA division involvement or include sufficient volume (estimate of >70ccs) of both superior/inferior or deep structures. All indeterminate images underwent secondary review (CJO). A random sampling of our 30 patients using an ABC/2 method [3] demonstrated that median volume was 122 cc. Below we outline the detailed process we applied to characterize each of the included radiographic features:

- *Stroke size*: $\geq$1/2 MCA territory was determined by visual estimate of a trained M.D. and underwent second review. Indeterminate images were reviewed for consensus by three authors.
- *Midline shift* was measured by a trained member of the team using imaging viewer software [Client Outlook, eUnity Diagnostic Viewer, version 6.10.2-489, for MacOs and PACS web viewer] by navigating to the level of the septum pellucidum at the slice of maximum MLS. The reviewer created a line connecting the attachment of falx cerebri anteriorly and the occipital protuberance. Windows were set at W:30 L:30. Distance between the midline and the septum pellucidum both the lateral and medial boundaries was measured by adding a line perpendicular to the midline. MLS used in the analysis was the average distance (mm) from the measurements of the lateral and medial boundaries. A blinded assessment of >10% of the data found the mean error was 0.19mm between the MLS reported in the radiographic reports and manual measurements.
- *Pineal gland shift* was measured at the level of the pineal gland, at the slice where the pineal gland shift was at its maximum. A line from the midline to the point of maximum and minimum deviation was taken, and averaged, to best obtain the deviation from the center of the pineal gland. Mean error between MD reviewers was 0.36mm.
- *ASPECTS*, or the Alberta Stroke Programme Early CT Score, divides the brain parenchyma into 10 separate non-overlapping regions, three regions at the level just rostral to the ganglionic structures and four at the level of the thalamus [4]. ASPECTS of 10 implies the brain parenchyma shows no evidence of ischemia, only at the two levels that the predetermined regions are examined. ASPECTS of 0 implies that all the predetermined areas have been affected by ischemia of the brain parenchyma, hypodensity, loss of cortical ribbon, and/or sulcal effacement. ASPECTS were generated using 5mm axial slices and set to variable window widths. The window, as described by Lev et al. [5], for optimal ischemic lesion identification is preferably set to W:30 L:30, or alternatively 40/40. As in Pexman et al. [6], patient positioning was reviewed to determine if the eyes were at the same level in the axial view. If >10% of each area had evidence of hypodensity or sulcal effacement, that area was marked as affected by ischemia and one point was deducted from the total score [4]. A review of 10% of scans showed a percent agreement between the two reviewers of 96.4% for dichotomous ASPECTS categories (10-8, 7-0), and 83.9% for high, medium, low ASPECTS (10-8, 7-4, 3-0). Cohen's Kappa was 0.647 for ASPECTS continuously.
- *Hemorrhage* was manually inspected according to ECASS II criteria. Petechial Hemorrhage was determined to be HI1 or HI2, and Parenchymal Hemorrhage PI1 or PI2 [7]. Percent agreement between trained team members (SC, CJO) on a 10% sample was 92.5%.
- *Cerebral atrophy* was recorded based on visual estimation of the ratio of sulcus to gyrus depth on the non-infarcted side and the ratio of the width of the caudate to the brain. A trained researcher evaluated admission non-contrast CT scans to determine the overall level of atrophy in the brain. Atrophy characterizations were categorized into no/mild atrophy (0) and moderate to severe atrophy (1). The percent agreement was 88%.
- *Basal Cistern Effacement* was determined (present/absent) in axial slices at the level of the orbitomeatal line after reviewing consecutive slices for evidence of partial or complete reduction of the basal cisterns space. Images were reviewed for consensus by three authors.

#### 2.4 Text-Derived Features Using Large Language Model Prediction

We applied transfer learning to fine-tune large language models from the pre-trained Clinical-Longformer model and derive text-driven predictions for each future MLS class and prediction window [8]. For the large language model text classifiers, we expanded the time window horizon to include the 36-hour task, in addition to the 8-hour and 24-hour horizons, to provide a more extensive view of the patient trajectory to the downstream models. These classifiers were trained using raw radiology report texts generated by clinicians at the time of each scan to describe the characteristics and diagnoses associated with a patient's stroke and edema state progression. In order to standardize the reports across hospitals, we cropped the texts to only include the "Findings" and "Impression" sections, which were available in reports from both datasets.

Before training, the texts were tokenized using the pre-trained Clinical-Longformer tokenizer [9]. Text data from the derivation set were split based on patient into training and test sets, with data from 80% patients being used for fine-tuning and 20% of patients being reserved for the testing set. By instantiating the pre-trained models as text classifiers, a final classification layer was added to the model head, taking the standard text prediction output of the model and transforming it to make soft-max probability predictions across the four defined MLS classes. We fine-tuned three separate text-classification large language models using the derivation dataset to predict the future maximum MLS value within windows of 8-hours, 24-hours, and 36-hours, respectively. The 36-hour horizon predictions provided more information about long-term MLS trajectory, but were not included for the downstream HELMET models as they were not deemed to be clinically relevant. The model's pre-trained weights were updated by minimizing the categorical cross-entropy loss between the predicted class probabilities and the true MLS class labels for each predictive window. The fine-tuning process included six epochs at an initial learning rate of $2*10^{-5}$, using the HuggingFace Transformers package in Python 3.11 and following previously established methods for the Clinical-Longformer model [8, 10]. Fine-tuning was conducted on a standard Microsoft Azure NC24ads A100 v4 virtual machine using a single Nvidia A100 GPU. After transfer learning was complete, the four class probabilities for each of the three large language models for each radiographic report were then incorporated into the datasets to be used as input features for the downstream ensemble learning models alongside the variables from radiographic images and patient medical records.

We report the modified AUROC metric for the multi-class classification task as defined in Section 4.4.1. Using derivation patient radiology reports, the large language model classifiers achieved AUROC scores of 72.23% for the 8-hour task, 67.25% for the 24-hour task, and 91.10% for the 36-hour task when tested on the derivation cohort. Applying the fine-tuned models to the external validation cohort, the large language model performance dropped to 48.04%, 50.13%, and 50.24% on the 8-hour, 24-hour, and 36-hour tasks respectively.

The output large language model class-based probabilities were then appended to the datasets. We forward-filled any missing hours prior to training of the downstream ensemble learning models.

#### 2.5 Missing Data & Frequency of Measurement

The derivation cohort was associated with a higher degree of missing data compared to the external validation cohort, especially in vital signs at admission and in collateral scores, first MLS value, HbA1c, and osmolality. The number and percentage of patients in both cohorts with missing information regarding the static variables are presented in Table 4. Our analysis also showed that vital signs were recorded more frequently at the Boston Medical Center, while laboratory data and radiographic variables were updated more frequently in the Massachusetts General Brigham cohort. These patterns may reflect differences in laboratory and scanning capacity across the two sites. For dynamic variables, the average time between updated measurements within the first week of hospitalization is presented across cohorts in Table 3.

### 3 Patient Cohorts

To further compare the relative severity of mass effect after infarct across sites, we present the breakdown of patients in each cohort by maximum MLS class reached over the course of their hospitalization in Table 2. No statistical significance was found between the proportion of patients in each maximum MLS class across the cohorts.

### 4 Machine Learning Model Design and Training

#### 4.1 Model Features and Feature Space Construction

We report a breakdown of each of the features used in our HELMET models. Table 5 describes the features used in the 24-hour prediction model while Table 6 describes the features used in the 8-hour prediction model. All date-time features are represented as number of hours after last seen well (e.g. if the patient was last seen well at 12:00 and the first MLS occurred at 18:00, the value of *firstmlsdt_time_censored* would be 6). Time censored variables are assigned a place holder value until the time of occurrence to prevent future information from being leaked into the present (e.g. if firstmlsdt_time_censored was 6, then the first five patient-hour observations would be set to -10).

#### 4.2 Comparison of XGBoost to Random Forest Models

We also tested Random Forest (RF) models [11] as part of our initial exploration (prior to adding the large language model-derived features). The Random Forest algorithm is an ensemble algorithm that creates multiple weak learner decision trees [11]. We once again assessed performance on the derivation dataset with five-fold cross-validation. XGBoost and RF achieved roughly equivalent performance, with a mean area under the receiver operating characteristic (AUROC) across the four classes of 0.86 (±0.017 for RF, ±0.0083 for XGBoost) for predicting 24-hour window maximum MLS, and 0.86 (±0.0066 for RF, ±0.0059 for XGBoost) for predicting 8-hour window maximum MLS. The relative similarity in the performance of XGBoost and RF models is consistent with their relatively similar ensemble tree architectures. In both settings, both models significantly outperformed the logistic regression baseline ($p<0.001$ on all tests for AUROC and AUPRC).

#### 4.3 Final Tuned Hyperparameters

Using a combination of cross-validation within runs and Bayesian updating across successive model training runs, we tuned the model hyperparameters and non-transition observation weights for both the HELMET-8 and HELMET-24 models. The final values for both models are presented in Table 8.

#### 4.4 Sensitivity Analyses

We conducted a number of sensitivity analyses to improve the performance of the model. First, we tested the inclusion of a sample weighting parameter which reduced the relative importance of samples where the target MLS class was the same as the patient's current MLS class (in order to further prioritize training on the more ambiguous cases of patient class transition). Results showed that a relative importance reduction of 0.4-0.7 yielded increased filtered performance at the expense of some overall, unfiltered performance. Next, we tested adding the class probabilities from the average transition kernel as features and adding the class probabilities as predicted by the linear regression model (discussed below). Neither of these feature additions led to any significant performance improvements. Finally, we tested reducing the feature space by removing the lowest-ranked features from the feature importance values of the initial model. Reducing the feature space to approximately 75 total features yielded increased performance, with further feature reductions being detrimental. However, any attempts at feature reduction proved detrimental after the addition of the large language model class predictions to the feature space.

### 5 Model Evaluation

#### 5.1 Complete Performance Metrics

In a filtered dataset using only prediction windows in which the patient's MLS class changed (the "filtered" task), the models result in a mean filtered AUROC of 94.8% (95% CI [93.0%, 96.6%]) for the 24-hour window and 80.6% (95% CI [66.7%, 94.5%]) for the 8-hour window on the derivation dataset. On the external validation dataset, the models result in a mean filtered AUROC of 57.59% (95% CI [55.4%, 59.8%]) for the 24-hour window and 63.5% (95% CI [54.9%, 72.1%]) for the 8-hour window. The filtered performance metrics show that the model performs well both when the patient is expected to remain in the same state and when the edema severity is changing. The complete unfiltered and filtered performance metrics, along with 95% confidence intervals are reported in Table 9.

#### 5.2 Receiver-Operator Characteristic Curves and Precision-Recall Curves

The receiver-operator characteristic (ROC) curves for both models and both patient cohorts are presented in Figure 1. Similarly, the precision-recall curves (PRC) for both models and both cohorts are presented in Figure 2.

#### 5.3 Shapley Additive Explanation Plots

SHapley Additive exPlanation (SHAP) values are derived from cooperative game theory, originally developed to fairly distribute payouts among players based on their contributions to an overall outcome. In the context of machine learning, SHAP values quantify the contribution of each feature to the prediction by simulating the model's behavior when each feature is included or excluded from the input data. Specifically, SHAP values represent the average marginal contribution of a feature across all possible combinations of feature subsets, ensuring that the contributions are fairly attributed in a manner consistent with the axioms of efficiency, symmetry, and additivity. SHAP plots indicate the aggregate influence of individual features on assigned classification categories. Each sub-bar's length indicates the mean SHAP value per class, highlighting the relative strength of a feature's influence on the prediction class outcome.

The features are organized by significance, with the most crucial ones positioned at the top. Figure 3 shows SHAP bar plots for the ensemble learning models.

### Supplementary Figures

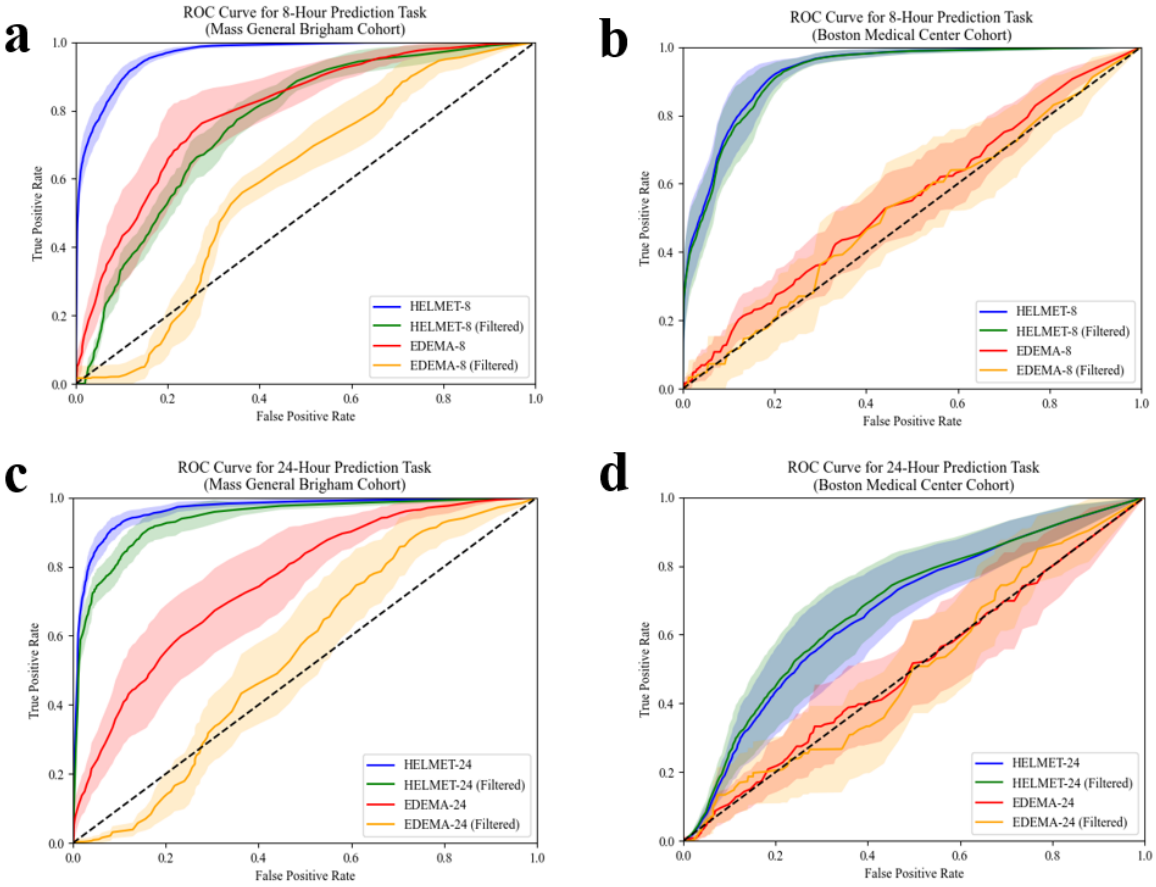


Figure 1: Receiver operating characteristic curves for HELMET models compared to baseline EDEMA Score models for overall and filtered datasets for both derivation and external validation cohorts. Blue lines show performance of the HELMET models on the overall dataset, Orange lines show performance of the HELMET models on the filtered (MLS transition) observations. Red and orange lines show performance of the baseline EDEMA Score models on the overall and filtered datasets, respectively. a) Performance on the derivation dataset for the 8-hour prediction task; b) performance on external validation dataset for the 8-hour prediction task; c) derivation cohort performance for 24-hour task; d) external validation cohort performance for 24-hour task

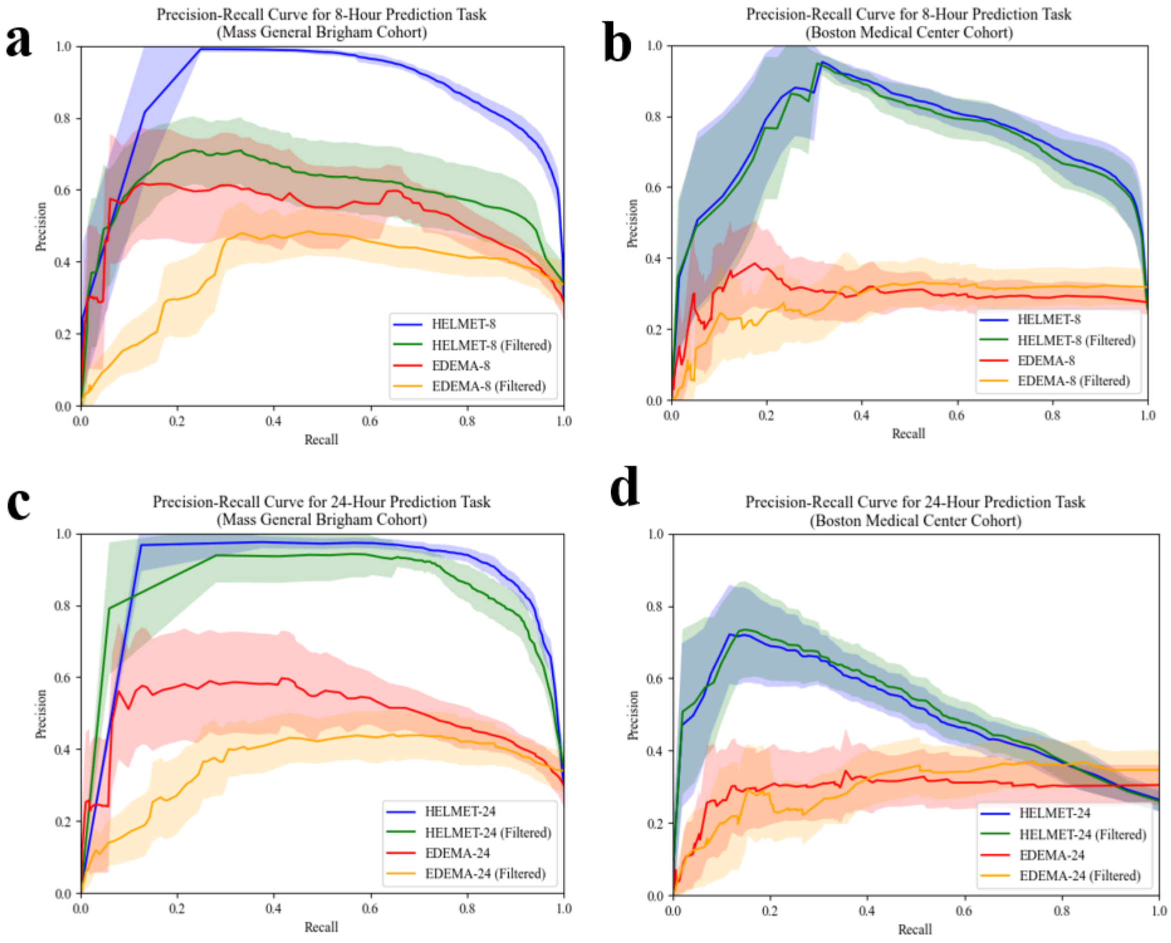


Figure 2: Precision recall curves for HELMET models compared to baseline EDEMA Score models for overall and filtered datasets for both derivation and external validation cohorts. Blue lines show performance of the HELMET models on the overall dataset, Orange lines show performance of the HELMET models on the filtered (MLS transition) observations. Red and orange lines show performance of the baseline EDEMA Score models on the overall and filtered datasets, respectively. a) Performance on the derivation dataset for the 8-hour prediction task; b) performance on external validation dataset for the 8-hour prediction task; c) derivation cohort performance for 24-hour task; d) external validation cohort performance for 24-hour task

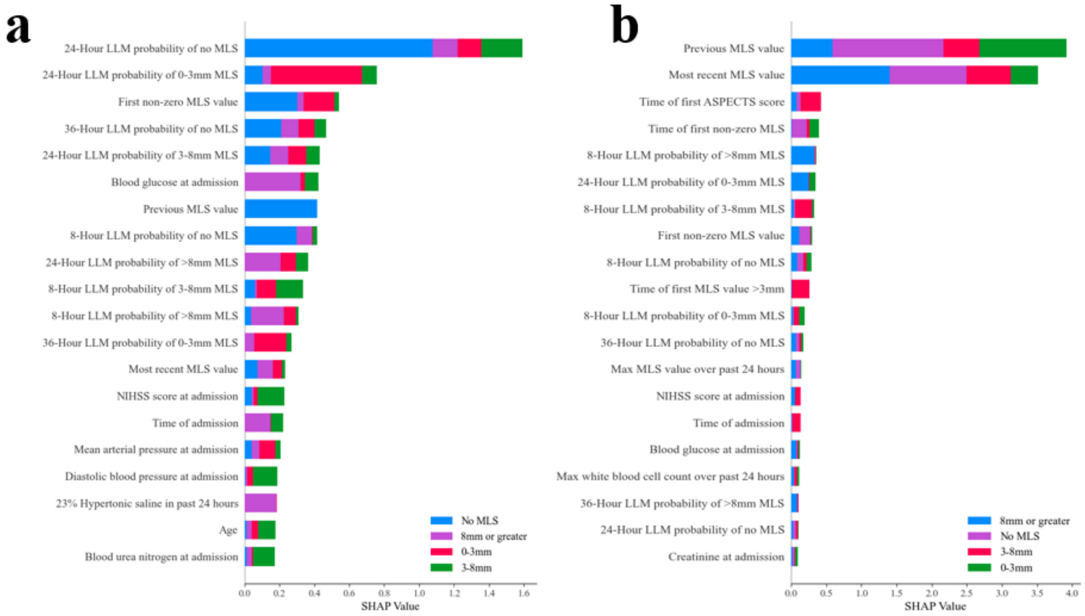


Figure 3: The 20 features with the highest SHAP values for the ensemble learning models. Colors represent contribution of each feature to positive predictions of each MLS class. a) Highest contributing features for the HELMET-24 b) Highest contributing features for HELMET-8.

### Supplementary Tables

Table 1: A summary of existing research on predicting events relating to cerebral edema.

| Study | Sample Size | Type | Prediction Target | AUROC | Dynamic Prediction? | Single Center? | Retrospective? | Other Limitations |
|---|---|---|---|---|---|---|---|---|
| Shimoyama et al., 2014 [12] | 119 | Logistic Regression | Malignant MCA infarction | 0.88 | No | Yes | Yes | Small sample biased towards an elderly population (average age 78.0) |
| Ong et al., 2017 [13] | 222 | Logistic Regression | Malignant brain edema | 0.75 | No | Yes | Yes | Late outcome prediction and only data from first 24 hours. |
| Cheng et al., 2020 [14] | 487 | Logistic Regression | Malignant brain edema | 0.8 | No | Yes | Yes | Same as Ong, et al., 2017. Also shows low generalizability without model re-training. |
| Wu et al., 2023 [15] | 1627 | Logistic Regression | Malignant brain edema | 0.9 | No | Yes | Yes | No imaging features used in the model. |
| Tang et al., 2024 [16] | 314 | Logistic Regression | Malignant brain edema | 0.88 | No | Yes | Yes | Homogeneous sample composition biased toward elderly population (average age 75.7) |

Table 2: Maximum MLS class reached by patients across cohorts and prediction tasks.

| Data set | Task | Maximum MLS class | | | |
|---|---|---|---|---|---|
| | | 0mm | 0-3mm | 3-8mm | >8mm |
| Mass General Brigham (derivation cohort) | 8-hour | 162 | 100 | 216 | 145 |
| | 24-hour | 123 | 81 | 204 | 125 |
| Boston Medical Center (validation cohort) | 8-hour | 10 | 10 | 17 | 23 |
| | 24-hour | 9 | 9 | 18 | 19 |

Table 3: Average time delay between updated measurements in dynamic variables within the first week of hospitalization across cohorts. All variables presented as mean hours (SD). Statistical significance of differences between cohorts tested using Mann-Whitney U test.

| Variable | Massachusetts General Brigham | Boston Medical Center | p-value |
|---|---|---|---|
| **Vital Signs** | | | |
| Heart rate | 6.1 (4.1) | 3.1 (4.2) | 0.000 |
| Systolic blood pressure | 7.1 (4.3) | 2.6 (3.8) | 0.000 |
| Diastolic blood pressure | 7.6 (4.7) | 3.1 (3.9) | 0.000 |
| Body temperature | 8.8 (6.5) | 5.9 (7.0) | 0.000 |
| **Laboratory Values** | | | |
| White blood cell count | 8.6 (7.2) | 13.8 (17.8) | 0.003 |
| Blood glucose | 6.0 (5.3) | 6.8 (7.6) | 0.652 |
| Osmolality | 11.8 (8.2) | 21.1 (23.5) | 0.008 |
| Creatinine | 7.8 (6.3) | 12.3 (12.8) | 0.002 |
| Sodium | 7.7 (6.2) | 10.2 (11.6) | 0.286 |
| Blood urea nitrogen | 8.2 (6.3) | 13.2 (15.6) | 0.003 |
| **Radiographic Measurements** | | | |
| Midline shift | 11.3 (9.1) | 18.8 (22.7) | 0.003 |
| Pineal gland shift | 11.6 (9.1) | 18.9 (22.4) | 0.008 |

Table 4: Missingness in static variables across cohorts. All variables presented as patient counts of missing data (percentage of patients). Statistical significance of differences between cohorts tested using $\chi^2$ test.

| Variable | Massachusetts General Brigham | Boston Medical Center | p-value |
|---|---|---|---|
| **Demographic Factors** | | | |
| Age | 0 (0%) | 0 (0%) | 1.000 |
| Sex | 0 (0%) | 0 (0%) | 1.000 |
| Race | 0 (0%) | 0 (0%) | 1.000 |
| **Medical History** | | | |
| Previous stroke | 0 (0%) | 0 (0%) | 1.000 |
| Atrial fibrillation | 0 (0%) | 0 (0%) | 1.000 |
| Hypertension | 0 (0%) | 0 (0%) | 1.000 |
| **Stroke Characteristics at Admission** | | | |
| NIHSS | 46 (7.4%) | 0 (0%) | 0.056 |
| ASPECTS | 0 (0%) | 0 (0%) | 1.000 |
| Stroke side | 0 (0%) | 0 (0%) | 1.000 |
| Anterior cerebral artery involved | 0 (0%) | 0 (0%) | 1.000 |
| Vessel Occlusion | 0 (0%) | 0 (0%) | 1.000 |
| Collateral Score | 207 (33.2%) | 28 (46.7%) | 0.051 |
| First MLS | 157 (25.2%) | 0 (0%) | 0.000 |
| Cerebral Atrophy | 3 (0.5%) | 0 (0%) | 1.000 |
| **Vital Signs at Admission** | | | |
| Mean arterial pressure | 43 (6.9%) | 0 (0%) | 0.068 |
| Systolic blood pressure | 0 (0%) | 0 (0%) | 1.000 |
| Diastolic blood pressure | 43 (6.9%) | 0 (0%) | 0.068 |
| Heart rate | 454 (72.9%) | 0 (0%) | 0.000 |
| Body temperature | 454 (72.9%) | 0 (0%) | 0.000 |
| **Laboratory Values at Admission** | | | |
| White blood cell count | 3 (0.5%) | 0 (0%) | 1.000 |
| Blood glucose | 2 (0.3%) | 0 (0%) | 1.000 |
| HbA1c | 98 (15.7%) | 3 (5.0%) | 0.041 |
| Osmolality | 445 (71.4%) | 31 (51.7%) | 0.002 |
| Creatinine | 3 (0.5%) | 0 (0%) | 1.000 |
| Sodium | 2 (0.3%) | 0 (0%) | 1.000 |
| Blood urea nitrogen | 3 (0.5%) | 0 (0%) | 1.000 |
| **Treatments Administered** | | | |
| Medical Thrombolysis | 0 (0%) | 0 (0%) | 1.000 |
| Mechanical Thrombectomy | 0 (0%) | 0 (0%) | 1.000 |
| Decompressive Hemicraniectomy | 0 (0%) | 0 (0%) | 1.000 |
| Osmotic Therapies | 0 (0%) | 0 (0%) | 1.000 |

Table 5: Complete list of features used by HELMET-24

| Feature | Relative Importance | Description | Source |
|---|---|---|---|
| LLM_24_3 | 53.3 | LLM-assessed probability of transition to class 3 over next 24 hours | Radiology Report Texts |
| LLM_24_2 | 46.8 | LLM-assessed probability of transition to class 2 over next 24 hours | Radiology Report Texts |
| LLM_24_1 | 41.4 | LLM-assessed probability of transition to class 1 over next 24 hours | Radiology Report Texts |
| LLM_24_0 | 29.1 | LLM-assessed probability of transition to class 0 over next 24 hours | Radiology Report Texts |
| LLM_36_1 | 11.5 | LLM-assessed probability of transition to class 1 over next 36 hours | Radiology Report Texts |
| wbc1 | 11.0 | White blood cell count at admission | Patient Medical Record |
| LLM_8_0 | 8.0 | LLM-assessed probability of transition to class 0 over next 8 hours | Radiology Report Texts |
| rolling_map | 7.8 | Maximum mean arterial pressure over past 24 hours | Patient Medical Record |
| LLM_8_2 | 7.5 | LLM-assessed probability of transition to class 2 over next 8 hours | Radiology Report Texts |
| firstmls_time_censored | 7.4 | Value of first measured MLS | Radiographic Images |
| rolling_pulse | 7.3 | Maximum pulse over past 24 hours | Patient Medical Record |
| LLM_36_0 | 6.9 | LLM-assessed probability of transition to class 0 over next 36 hours | Radiology Report Texts |
| mls5dt_time_censored | 6.8 | Time at which patient first reached MLS of 5mm or greater | Radiographic Images |
| temp | 6.7 | Most recent body temperature | Patient Medical Record |
| prev_mls | 6.5 | Last measured MLS value prior to current value | Radiographic Images |
| LLM_36_2 | 6.4 | LLM-assessed probability of transition to class 2 over next 36 hours | Radiology Report Texts |
| LLM_8_3 | 5.9 | LLM-assessed probability of transition to class 3 over next 8 hours | Radiology Report Texts |
| gluc1 | 5.7 | Blood glucose at admission | Patient Medical Record |
| hi2 | 5.6 | Petechial hemorrhage gaining confluence in most recent scan | Radiographic Images |
| rolling_size_mls | 5.6 | Maximum MLS value over past 24 hours | Radiographic Images |
| pulse1 | 5.1 | Pulse at admission | Patient Medical Record |
| size_mls | 4.9 | Most recent MLS value | Radiographic Images |
| rolling_temp | 4.8 | Maximum body temperature over past 24 hours | Patient Medical Record |
| pgs2_time_censored | 4.7 | Value of maximum pineal gland shift of 2mm or greater so far | Radiographic Images |
| pgs4dt_time_censored | 4.7 | Time at which pineal gland shift first reached 4mm or greater | Radiographic Images |
| temp1 | 4.7 | Body temperature at admission | Patient Medical Record |
| hts23_x | 4.5 | Administration of hypertonic saline (concentration 23.4%) | Patient Medical Record |
| evd | 4.5 | Extraventricular drain in most recent scan | Radiographic Images |
| aspects | 4.4 | Most recent ASPECTS index score | Radiographic Images |
| rolling_hts3_y | 4.1 | Whether hypertonic saline (concentration 3%) has been administered | Patient Medical Record |
| aspects1dt_time_censored | 4.1 | Time of first ASPECTS index score | Radiographic Images |
| pres | 4.0 | Time of admission | Patient Medical Record |
| map1 | 4.0 | Mean arterial pressure at admission | Patient Medical Record |
| LLM_36_3 | 4.0 | LLM-assessed probability of transition to class 3 over next 36 hours | Radiology Report Texts |
| bun1 | 4.0 | Blood urea nitrogen at admission | Patient Medical Record |
| wbc | 4.0 | Most recent white blood cell count | Patient Medical Record |
| imagetype | 3.8 | Categorical representation of most recent scan type | Radiographic Images |
| rolling_aca_x | 3.7 | Whether anterior cerebral artery has been involved in past 24 hours | Radiographic Images |

Continued on next page

**Table 5 – continued from previous page**

| Feature | Relative Importance | Description Source | |
|---|---|---|---|
| rolling_hts23_y | 3.7 | Whether hypertonic saline (concentration 23.4%) has been administered | Patient Medical Record |
| firstmlsdt_time_censored | 3.5 | Time of first MLS measurement greater than 0 | Radiographic Images |
| stroke_territory | 3.5 | Size of affected stroke territory | Radiographic Images |
| rolling_osm | 3.5 | Maximum osmolality value over past 24 hours | Patient Medical Record |
| mls3dt_time_censored | 3.5 | Time of first MLS measurement of 3mm or greater | Radiographic Images |
| sbp1 | 3.5 | Systolic blood pressure at admission | Patient Medical Record |
| rolling_size_pgs | 3.4 | Maximum pineal gland shift over past 24 hours | Radiographic Images |
| rolling_gluc | 3.3 | Maximum blood glucose over past 24 hours | Patient Medical Record |
| rolling_sbp | 3.3 | Maximum systolic blood pressure over past 24 hours | Patient Medical Record |
| sbp | 3.3 | Most recent systolic blood pressure | Patient Medical Record |
| age_calc | 3.3 | Age | Patient Medical Record |
| cr | 3.3 | Most recent creatinine value | Patient Medical Record |
| dbp1 | 3.2 | Diastolic blood pressure at admission | Patient Medical Record |
| rolling_dbp | 3.2 | Maximum diastolic blood pressure over past 24 hours | Patient Medical Record |
| rolling_wbc | 3.1 | Maximum white blood cell count over past 24 hours | Patient Medical Record |
| LLM_8_1 | 3.1 | LLM-assessed probability of transition to class 1 over next 8 hours | Radiology Report Texts |
| aspects1_time_censored | 3.0 | ASPECTS score from first scan | Radiographic Images |
| osm | 3.0 | Most recent osmolality value | Patient Medical Record |
| mtdt_time_censored | 3.0 | Time of mechanical thrombectomy | Patient Medical Record |
| aca_x | 2.9 | Whether anterior cerebral artery is involved in most recent scan | Radiographic Images |
| htn | 2.9 | History of hypertension | Patient Medical Record |
| rolling_mannitol_y | 2.9 | Whether mannitol has been administered | Patient Medical Record |
| stroke | 2.9 | Occurrence of previous stroke | Patient Medical Record |
| rolling_na | 2.8 | Maximum sodium over past 24 hours | Patient Medical Record |
| dbp | 2.8 | Most recent diastolic blood pressure value | Patient Medical Record |
| af | 2.7 | History of atrial fibrillation | Patient Medical Record |
| na1 | 2.7 | Sodium at admission | Patient Medical Record |
| mt_time_censored | 2.7 | Whether mechanical thrombectomy has occurred | Patient Medical Record |
| dt | 2.7 | Hours since Last Seen Well | Patient Medical Record |
| rolling_bun | 2.6 | Maximum blood urea nitrogen in past 24 hours | Patient Medical Record |
| rolling_imagetype | 2.6 | Most intensive scan type in last 24 hours | Radiographic Images |
| rolling_aspects | 2.6 | Maximum ASPECTS score in past 24 hours | Radiographic Images |
| bun | 2.6 | Most recent blood urea nitrogen value | Patient Medical Record |
| nihss | 2.5 | NIHSS score at admission | Patient Medical Record |
| sex | 2.5 | Female or not | Patient Medical Record |
| tpa_time_censored | 2.5 | Administration of TPA | Patient Medical Record |
| rolling_hts3_x | 2.4 | Administration of hypertonic saline (concentration 3%) in past 24 hours | Patient Medical Record |
| size_pgs | 2.4 | Most recent pineal gland shift measurement | Radiographic Images |

Continued on next page

**Table 5 – continued from previous page**

| Feature | Relative Importance | Description Source | |
|---|---|---|---|
| hts3_x | 2.4 | Administration of hypertonic saline (concentration 3%) in current hour | Patient Medical Record |
| tpadt_time_censored | 2.4 | Time at which TPA was administered | Patient Medical Record |
| rolling_cr | 2.4 | Maximum creatinine value in past 24 hours | Patient Medical Record |
| rolling_osmotics_y | 2.4 | Whether osmotic treatment has been administered | Patient Medical Record |
| mls12dt_time_censored | 2.4 | Time at which MLS first reached 12mm or greater | Radiographic Images |
| mannitol_x | 2.3 | Administration of mannitol in current hour | Patient Medical Record |
| gluc | 2.3 | Most recent blood glucose value | Patient Medical Record |
| na | 2.3 | Most recent sodium value | Patient Medical Record |
| rolling_hts23_x | 2.2 | Administration of hypertonic saline (concentration 23.4%) in past 24 hours | Patient Medical Record |
| obsTime | 2.2 | Current date and time | |
| cr1 | 2.2 | Creatinine at admission | Patient Medical Record |
| rolling_hi1 | 2.1 | Petechial hemorrhage in past 24 hours | Radiographic Images |
| rolling_stroke_territory | 2.0 | Maximum size of stroke territory affected in last 24 hours | Radiographic Images |
| rolling_bce_severity | 2.0 | Maximum basilar cistern effacement in past 24 hours | Radiographic Images |
| pgs2dt_time_censored | 1.9 | Time at which pineal gland shift first reached 2mm or greater | Radiographic Images |
| rolling_hi2 | 1.9 | Petechial hemorrhages starting to gain confluence in past 24 hours | Radiographic Images |
| map | 1.9 | Most recent mean arterial pressure value | Patient Medical Record |
| hi1 | 1.8 | Petechial hemorrhage present in most recent scan | Radiographic Images |
| rolling_aca_y | 1.8 | Whether anterior cerebral artery was ever involved | Radiographic Images |
| rolling_evd | 1.6 | Extraventricular drain in past 24 hours | Radiographic Images |
| pulse | 1.6 | Most recent pulse | Patient Medical Record |
| rolling_mannitol_x | 1.5 | Administration of mannitol in past 24 hours | Patient Medical Record |

Table 6: Complete list of features used by HELMET-8

| Feature | Relative Importance | Description | Source |
|---|---|---|---|
| prev_mls | 72.7 | Last measured MLS value prior to current value | |
| LLM_8_2 | 48.9 | LLM-assessed probability of transition to class 2 over next 8 hours | |
| size_mls | 47.1 | Most recent MLS measurement | |
| LLM_8_3 | 42.0 | LLM-assessed probability of transition to class 3 over next 8 hours | |
| LLM_8_1 | 37.4 | LLM-assessed probability of transition to class 1 over next 8 hours | |
| firstmlsdt_time_censored | 14.0 | Time of first non-zero MLS measurement | |
| mls3dt_time_censored | 13.6 | Time of first MLS measurement greater than 3mm | |
| LLM_8_0 | 12.5 | LLM-assessed probability of transition to class 0 over next 8 hours | |
| firstmls_time_censored | 10.7 | Value of first non-zero MLS measurement | |
| pulse1 | 9.6 | Pulse at admission | |
| temp1 | 9.4 | Body temperature at admission | |
| rolling_hts3_x | 8.5 | Whether hypertonic saline (concentration 3%) has been administered in past 24 hours | |
| LLM_24_3 | 8.2 | LLM-assessed probability of transition to class 3 over next 24 hours | |
| LLM_24_1 | 7.8 | LLM-assessed probability of transition to class 1 over next 24 hours | |
| dbp | 7.5 | Most recent diastolic blood pressure | |
| rolling_temp | 7.1 | Maximum body temperature in past 24 hours | |
| LLM_24_0 | 6.9 | LLM-assessed probability of transition to class 0 over next 24 hours | |
| pgs2dt_time_censored | 6.8 | Time of first pineal gland shift measurement greater than 2mm | |
| LLM_36_3 | 6.5 | LLM-assessed probability of transition to class 3 over next 36 hours | |
| aspects1dt_time_censored | 6.3 | Time of first ASPECTS score | |
| pulse | 6.0 | Most recent pulse | |
| LLM_24_2 | 5.9 | LLM-assessed probability of transition to class 2 over next 24 hours | |
| imagetype | 5.9 | Category of most recent scan | |
| pres | 5.9 | Time of admission | |
| LLM_36_0 | 5.7 | LLM-assessed probability of transition to class 0 over next 36 hours | |
| rolling_dbp | 5.6 | Maximum diastolic blood pressure in past 24 hours | |
| LLM_36_2 | 5.5 | LLM-assessed probability of transition to class 2 over next 36 hours | |
| rolling_imagetype | 5.4 | Most intense scan in past 24 hours | |
| af | 5.4 | History of atrial fibrillation | |
| LLM_36_1 | 5.3 | LLM-assessed probability of transition to class 1 over next 36 hours | |
| mt_time_censored | 5.3 | Whether mechanical thrombectomy has occurred | |
| pgs4dt_time_censored | 5.1 | Time of first pineal gland shift measurement greater than 4mm | |
| rolling_osmotics_y | 5.1 | Whether osmotics have been administered | |
| rolling_sbp | 4.9 | Maximum systolic blood pressure in past 24 hours | |
| hts23_x | 4.8 | Administration of hypertonic saline (concentration 23.4%) in current hour | |
| tpa_time_censored | 4.6 | Whether TPA has been administered | |
| rolling_aspects | 4.6 | Maximum ASPECTS score in past 24 hours | |
| stroke_territory | 4.5 | Size of affected stroke territory on most recent scan | |

Continued on next page

**Table 6 – continued from previous page**

| Feature | Relative Importance | Description | Source |
|---|---|---|---|
| rolling_aca_x | 4.4 | Whether anterior cerebral artery has been affected in past 24 hours | |
| cr1 | 4.4 | Creatine at admission | |
| wbc | 4.4 | Most recent white blood cell count | |
| rolling_hts23_x | 4.4 | Whether hypertonic saline (concentration 23.4%) has been administered in past 24 hours | |
| rolling_map | 4.3 | Maximum mean arterial pressure in past 24 hours | |
| rolling_pulse | 4.2 | Maximum pulse in past 24 hours | |
| rolling_size_pgs | 4.1 | Maximum pineal gland shift measurement in past 24 hours | |
| rolling_size_mls | 4.1 | Maximum MLS measurement in past 24 hours | |
| wbc1 | 4.1 | White blood cell count at admission | |
| aspects1_time_censored | 4.1 | ASPECTS score at admission | |
| bun | 4.1 | Most recent blood urea nitrogen value | |
| dbp1 | 4.1 | Diastolic blood pressure at admission | |
| size_pgs | 4.0 | Most recent pineal gland shift measurement | |
| rolling_wbc | 4.0 | Maximum white blood cell count in past 24 hours | |
| mtdt_time_censored | 4.0 | Time of mechanical thrombectomy | |
| cr | 4.0 | Most recent creatinine value | |
| na | 4.0 | Most recent sodium value | |
| obsTime | 3.9 | Current time | |
| map1 | 3.9 | Mean arterial pressure at admission | |
| aca_x | 3.9 | Whether anterior cerebral artery was involved in most recent scan | |
| rolling_stroke_territory | 3.8 | Maximum size of affected stroke territory in past 24 hours | |
| rolling_aca_y | 3.8 | Whether anterior cerebral artery has been involved in scans in the past 24 hours | |
| rolling_gluc | 3.8 | Maximum blood glucose in past 24 hours | |
| gluc1 | 3.7 | Blood glucose at admission | |
| temp | 3.7 | Most recent body temperature | |
| map | 3.7 | Most recent mean arterial pressure | |
| tpadt_time_censored | 3.7 | Time at which TPA was administered | |
| bun1 | 3.6 | Blood urea nitrogen at admission | |
| aca1 | 3.5 | Whether anterior cerebral artery was involved at first scan | |
| rolling_mannitol_x | 3.5 | Whether mannitol has been administered in last 24 hours | |
| nihss | 3.5 | NIHSS score at admission | |
| age_calc | 3.5 | Age | |
| na1 | 3.4 | Sodium at admission | |
| sbp | 3.4 | Most recent systolic blood pressure | |
| dt | 3.4 | Current time | |
| rolling_mannitol_y | 3.4 | Whether mannitol has been administered at any point | |
| rolling_cr | 3.2 | Maximum creatinine value in past 24 hours | |
| mannitol_x | 3.1 | Administration of mannitol | |

Continued on next page

**Table 6 – continued from previous page**

| Feature | Relative Importance | Description | Source |
|---|---|---|---|
| sbp1 | 3.1 | Systolic blood pressure at admission | |
| aspects | 3.0 | Most recent ASPECTS score | |
| rolling_na | 2.9 | Maximum sodium value in past 24 hours | |
| sex | 2.8 | Female or not | |
| osm | 2.6 | Most recent osmolality value | |
| gluc | 2.6 | Most recent blood glucose value | |
| rolling_bce_severity | 2.5 | Maximum basilar cistern effacement in past 24 hours | |
| rolling_osm | 2.4 | Maximum osmolality in past 24 hours | |
| rolling_bun | 2.4 | Maximum blood urea nitrogen in past 24 hours | |
| rolling_hts3_y | 2.3 | Whether hypertonic saline (concentration 3%) has been administered | |
| hi2 | 2.2 | Petechial hemorrhage on most recent scan | |
| mls7_time_censored | 2.0 | Value of maximum MLS measurement greater than 7mm | |
| mls7dt_time_censored | 1.8 | Time of first MLS measurement greater than 7mm | |
| pgs4_time_censored | 1.8 | Value of maximum pineal gland shift greater than 4mm | |
| mls5dt_time_censored | 1.7 | Time of first MLS measurement greater than 5mm | |
| rolling_hts23_y | 1.6 | Whether hypertonic saline (concentration 23.4%) has been administered | |
| hts3_x | 1.6 | Administration of hypertonic saline (concentration 3%) | |
| rolling_hi1 | 1.1 | Petechial hemorrhage in past 24 hours | |
| rolling_hi2 | 0.1 | Petechial hemorrhage gaining confluence in past 24 hours | |

Table 7: A comparison of XGBoost and Random Forest performance on five-fold splits on the derivation dataset.

| Prediction Task | Model | AUROC | AUPRC | Accuracy |
|---|---|---|---|---|
| 24 hours | Random Forrest | 86.0%(84.51%, 87.49%) | 68.0%(64.41%, 71.59%) | 62.0%(58.32%, 65.68%) |
| | XGBoost | 86.1%(85.37%, 86.83%) | 68.7%(67.87%, 69.53%) | 62.1%(60.53%, 63.67%) |
| 8 hours | Random Forrest | 86.0%(85.42%, 86.58%) | 67.0%(65.33%, 68.67%) | 63.0%(59.14%, 66.86%) |
| | XGBoost | 86.2%(85.68%, 86.72%) | 67.3%(66.84%, 67.76%) | 62.8%(61.61%, 63.99%) |

Table 8: Final hyperparameters used in training HELMET-24 and HELMET-8. Optimal hyperparameters were determined through cross-validation and bayesian optimizaiton across successive model training runs.

| Parameter | HELMET-24 | HELMET-8 |
|---|---|---|
| Maximum Tree Depth | 7 | 4 |
| Minimum Child Weight | 1 | 10 |
| Learning Rate | 0.0972 | 0.0669 |
| Gamma | 1.32 | 0.0831 |
| Data Subsample | 0.704 | 0.768 |
| Regularization Lambda | 6.07 | 6.62 |
| Regularization Alpha | 2.04 | 1.36 |
| No-transition Sample Weight | 0.475 | 0.563 |

Table 9: Comparison of performance metrics for HELMET and EDEMA models across derivation and external validation cohorts. Reported values were averaged across five cross-validation folds. The HELMET models were derived using the training set of the derivation cohort and were evaluated on the testing set of the derivation cohort and the entire external validation cohort. The EDEMA Regression Baseline models were separately trained and tested on both the derivation and external validation data. Values in parentheses reflect the 95% confidence intervals.

| Cohort | Metric | Type | 24 hours ahead | | 8 hours ahead | |
|---|---|---|---|---|---|---|
| | | | **EDEMA Baseline** | **HELMET-24** | **EDEMA Baseline** | **HELMET-8** |
| **Derivation Cohort** | AUROC | Overall | 77.7% (71.0%, 84.5%) | 96.7% (95.2%, 98.1%) | 80.5% (74.1%, 86.9%) | 96.6% (95.6%, 97.7%) |
| | | Filtered | 54.9% (41.3%, 68.5%) | 94.1% (92.3%, 96.0%) | 59.6% (47.2%, 71.9%) | 76.2% (66.6%, 85.8%) |
| | AUPRC | Overall | 57.1% (41.2%, 73.0%) | 87.2% (83.2%, 91.2%) | 59.3% (44.9%, 73.6%) | 87.5% (82.9%, 92.1%) |
| | | Filtered | 37.1% (29.4%, 44.9%) | 85.2% (78.1%, 92.2%) | 40.9% (32.4%, 49.4%) | 61.2% (46.9%, 75.5%) |
| | Accuracy | Overall | 56.2% (49.2%, 63.2%) | 87.3% (84.1%, 90.4%) | 62.0% (57.7%, 66.3%) | 82.9% (76.2%, 89.6%) |
| | | Filtered | 24.8% (20.7%, 29.0%) | 81.2% (76.5%, 85.9%) | 24.8% (18.2%, 31.4%) | 46.7% (38.1%, 55.3%) |
| | Sensitivity | Overall | 40.3% (31.4%, 49.1%) | 91.2% (85.4%, 97.0%) | 34.3% (29.1%, 39.4%) | 57.7% (47.4%, 68.1%) |
| | | Filtered | 40.3% (31.4%, 49.1%) | 91.2% (85.4%, 97.0%) | 34.3% (29.1%, 39.4%) | 57.7% (47.4%, 68.1%) |
| | Specificity | Overall | 80.6% (66.8%, 94.3%) | 94.0% (88.5%, 99.5%) | 87.1% (78.1%, 96.1%) | 90.2% (81.1%, 99.2%) |
| | | Filtered | 88.6% (74.7%, 100.0%) | 97.8% (91.1%, 100.0%) | 86.3% (64.2%, 100.0%) | 75.6% (9.5%, 100.0%) |
| **External Validation Cohort** | AUROC | Overall | 49.0% (37.6%, 60.4%) | 69.7% (60.3%, 79.0%) | 55.3% (46.6%, 64.0%) | 92.5% (89.4%, 95.6%) |
| | | Filtered | 48.0% (32.3%, 63.7%) | 70.7% (61.1%, 80.3%) | 51.8% (41.8%, 61.8%) | 92.1% (88.5%, 95.7%) |
| | AUPRC | Overall | 31.7% (19.3%, 44.1%) | 46.9% (35.6%, 58.1%) | 34.1% (23.1%, 45.1%) | 80.5% (69.8%, 91.3%) |
| | | Filtered | 34.2% (23.2%, 45.3%) | 48.4% (36.2%, 60.6%) | 33.2% (22.5%, 43.9%) | 79.8% (70.3%, 89.3%) |
| | Accuracy | Overall | 31.9% (21.5%, 42.2%) | 48.6% (46.7%, 50.6%) | 31.9% (16.7%, 47.2%) | 75.3% (70.3%, 80.3%) |
| | | Filtered | 27.5% (13.1%, 41.8%) | 48.7% (45.6%, 51.8%) | 22.7% (5.6%, 39.8%) | 73.3% (68.0%, 78.6%) |
| | Sensitivity | Overall | 69.5% (47.8%, 91.2%) | 87.4% (80.9%, 94.0%) | 55.4% (35.5%, 75.3%) | 92.1% (89.4%, 94.9%) |
| | | Filtered | 69.5% (47.8%, 91.2%) | 87.4% (80.9%, 94.0%) | 55.4% (35.5%, 75.3%) | 92.1% (89.4%, 94.9%) |
| | Specificity | Overall | 64.5% (37.5%, 91.5%) | 80.6% (75.3%, 85.9%) | 69.5% (62.2%, 76.8%) | 94.1% (89.6%, 98.6%) |
| | | Filtered | 85.0% (44.5%, 100.0%) | 95.0% (91.5%, 98.5%) | 82.1% (64.9%, 99.3%) | 99.2% (96.8%, 100.0%) |
